# Diffusion-based vertex-wise connectomics encodes individual differences in cognition, affect and pathology

**DOI:** 10.64898/2026.11.18.26368444

**Authors:** Jose Pineda, María Cabello-Toscano, Elisabet Sarri, Saül Pascual-Diaz, the Alzheimer’s Disease Neuroimaging Initiative

**Affiliations:** Department of Medicine, University of Barcelona, Barcelona, Spain; Faculty of Medicine, University of Vic - Central University of Catalonia (UVic-UCC), Vic, Spain; Tissue Repair and Regeneration Laboratory (TR2Lab), Institute for Research and Innovation in Life and Health Sciences in Central Catalonia (IrisCC), Vic, Spain; Leenaards Memory Centre, Department of Clinical Neurosciences, Lausanne University Hospital and University of Lausanne, Lausanne, Switzerland; Institute of Neuroscience, University of Barcelona, Barcelona, Spain; IDIBAPS, Institut d’Investigacions Biomèdiques August Pi i Sunyer, Barcelona, Spain; Center for Neuroscience Imaging Research, Institute for Basic Science (IBS), Suwon, South Korea

## Abstract

No two brains are wired alike, and much of a person’s cognitive and clinical state is written in this individual variation, not in the group average. Structural connectomes, however, are typically read through group atlases that summarize connectivity into regional averages, leaving this individual signal untapped. We build a vertex-wise structural connectome, in which every cortical surface vertex is a node, and apply it across the Alzheimer’s continuum. Using an independent dataset for methodological development and a multisite cohort of 317 participants from ADNI-4, we show that connection distance and organizational scale form a nested hierarchy, from a long-range backbone to fine, short-range structure. The individual fingerprint is strongest in medium- and short-range connections and becomes less distinctive as connectivity is pooled into atlas parcels. This individual structure can be recovered across spatial scales, although its fidelity depends on diffusion acquisition quality, with multishell data providing a more informative reference than single-shell data. In the aging brain, an individually referenced balance between short- and long-range connectivity is associated with episodic memory, cognitive reserve, depressive symptoms, and shared molecular pathology. While cognitive associations largely reflect diagnostic differences, associations with depressive symptoms and pathology burden remain detectable after adjustment for diagnosis. These findings show that individual structural connectomes retain clinically and biologically relevant variation that is not fully captured by diagnostic classification or group-level summaries.

## Introduction

Human brain organization varies substantially across individuals, and this variability is increasingly recognized not as noise around a canonical brain but as a biologically and clinically meaningful signal in its own right (Mueller et al. 2013; Mansour Let al. 2021). Most neuroimaging, however, characterizes the brain by comparing group means within predefined atlas parcels, an approach that describes the average brain while discarding the subject-specific features that may be the ones of clinical interest.

A subject-specific framework has been proposed for functional MRI to recover this lost information, characterizing each brain densely enough to map its own organization rather than pooling participants (Laumann et al. 2015; Braga and Buckner 2017; Gordon et al. 2017, 2026). An equivalent framework has not been established for structural connectivity, which is still analyzed almost entirely through group atlases and regional averages. Yet the raw material for one is clearly present. Whole-brain white-matter architecture is distinctive enough to identify a person from diffusion MRI with near-perfect accuracy (Yeh et al. 2016), an identifiability that can be quantified as a fingerprint score (Amico and Goñi 2018), that strengthens with spatial resolution, and that concentrates in heteromodal association cortex (Mansour L et al. 2021; Mueller et al. 2013).

In parallel, the connectome is organized as a multiscale, hierarchically nested community structure (Betzel and Bassett 2017) whose form is strongly shaped by the physical cost of connection distance, to the point of self-similarity across scales (Zheng et al. 2020). These two literatures have largely developed apart, and at coarse spatial grain. Identifiability has been measured mostly at the voxel (Yeh et al. 2016) or atlas-parcel (Amico and Goñi 2018) level, while multiscale community structure has been characterized on regional graphs rather than at the resolution of the cortical surface itself. As a result, three questions remain open: whether connection distance and community scale are distinct organizational axes or a single fused one; whether the individual fingerprint is carried preferentially by connections of a particular length; and whether the structure that distinguishes individuals is preserved under atlas parcellation, and at which spatial scale it is most informative.

Alzheimer’s disease (AD), the leading cause of dementia in later life, offers a natural test of whether this individual structure matters. AD is defined biologically by amyloid and tau pathology (Jack et al. 2018), yet that pathology is not distributed uniformly across patients. Tau deposition follows distinct spatiotemporal trajectories in different individuals, and these trajectories map onto the clinical variants of the disease: alongside the common amnestic form are presentations that begin with visual, language, executive, or behavioral failure, each tracking the regional distribution of tau and neurodegeneration more closely than the more diffuse burden of amyloid (Vogel et al. 2021; Jones et al. 2024). This heterogeneity is not independent of the brain’s wiring: models in which pathology spreads along brain networks reproduce the spatiotemporal evolution of tau across the cortex (Vogel et al. 2021; Raj et al. 2025). If the relationship between pathology and connectivity is spatially specific and differs between individuals, an individually resolved, vertex-wise structural connectome should capture disease-relevant information that regional and group-level representations discard.

The same heterogeneity extends to how individuals carry the disease. Two axes are central here: individuals with similar levels of pathology differ markedly in the cognition they retain, a capacity termed cognitive reserve (Stern et al. 2020; Pappalettera et al. 2024), and in their affective burden, with depression bidirectionally entwined with the disease as risk factor, prodrome, and symptom (Huang et al. 2024). Whether an individually resolved structural connectome carries information about these dimensions beyond what diagnostic classification already captures is an important clinical question.

Here we develop a vertex-wise structural connectome, in which every cortical surface vertex is a node, and show that it retains individual-specific structural information that atlas parcellation discards. To make this approach accessible and reproducible, we provide an open, interactive toolbox that guides users through the construction and analysis of vertex-wise structural connectomes and generates scripts for application to their own data. Where vertex-level connectivity representations have been recently used to improve cross-subject correspondence (Zhou et al. 2026), we characterize each connectome individually, without aligning its organization to a group representation. We find that connection distance and organizational scale are coupled in a nested hierarchy that is reproducible within individuals and distinctive between them. This subject-specific pattern is carried most distinctively by medium- and short-range connections. Although individuals remain identifiable across spatial scales, vertex-wise representation retains more individual-specific information than atlas representations. Finally, we relate an individually referenced short-to-long connectivity balance to clinical and pathological dimensions across the Alzheimer’s continuum. Although its associations with episodic memory and cognitive reserve largely reflect diagnostic differences, associations with depressive symptoms and shared molecular pathology remain detectable after controlling for diagnosis, highlighting individual variation that is not fully captured by diagnostic classification.

We establish these findings in two independent cohorts: a dataset used for methodological development and acquisition-related analyses (Aerts et al. 2018), and a multisite ADNI cohort comprising 317 participants across the Alzheimer’s continuum (Jack et al. 2008). The individual structural connectome thus emerges not as a refinement of the group average but as a distinct level of description, one that complements group-level analyses, that carries clinically meaningful information about the person, and that requires individual data of sufficient quality to recover.

## Results

### Demographics

We developed the method on the BTC dataset (OpenNeuro ds001226; Aerts et al. 2022, 2018), a multi-shell diffusion-MRI dataset, using 10 healthy control subjects (4 women; aged 48–75 years; mean 59.6, SD = 10.3; one further control was excluded at quality control, Supplementary Materials S1) to assess the conditions under which subject-specific structure can be recovered. We then applied the validated method to 322 ADNI-4 participants spanning the Alzheimer’s continuum (Weiner et al. 2023): 218 cognitively normal (CN), 80 with mild cognitive impairment (MCI), 23 with AD, and a single non-AD dementia case. Before the analysis, we defined two inclusion thresholds based on connectome-construction diagnostics, selected by inspecting their cohort distributions as indicators of potentially inadequate T1-to-diffusion registration: streamline endpoints had to reach at least 40% of white-surface vertices and lie a median of no more than 5mm from the surface. Four participants failed these criteria (one CN, one MCI, and two AD) and were excluded from all analyses, as was the single non-AD case, which lies outside the Alzheimer’s continuum, leaving 317 participants: 217 CN, 79 MCI, and 21 AD (Fig. 1; participant flow in Supplementary Materials S1). Age differed across diagnostic groups (CN 70.2, MCI 73.7, AD 75.9 years, one-way ANOVA *p* < .001), education differed and was lowest in AD (CN 16.5, MCI 16.0, AD 15.1 years; *p* = .035) and the proportion of women was higher in CN (67%) than MCI (48%), with AD intermediate (58%; χ^2^ *p* = .011). Age was unavailable for 16 participants (eight of whom also lacked sex); these participants were omitted from analyses requiring age and sex as covariates, which therefore used 301 participants (208 CN, 74 MCI, and 19 AD) from 19 sites. Accordingly, subsequent analyses accounted for differences in age, sex, and education across diagnostic groups.

**Figure 1.**
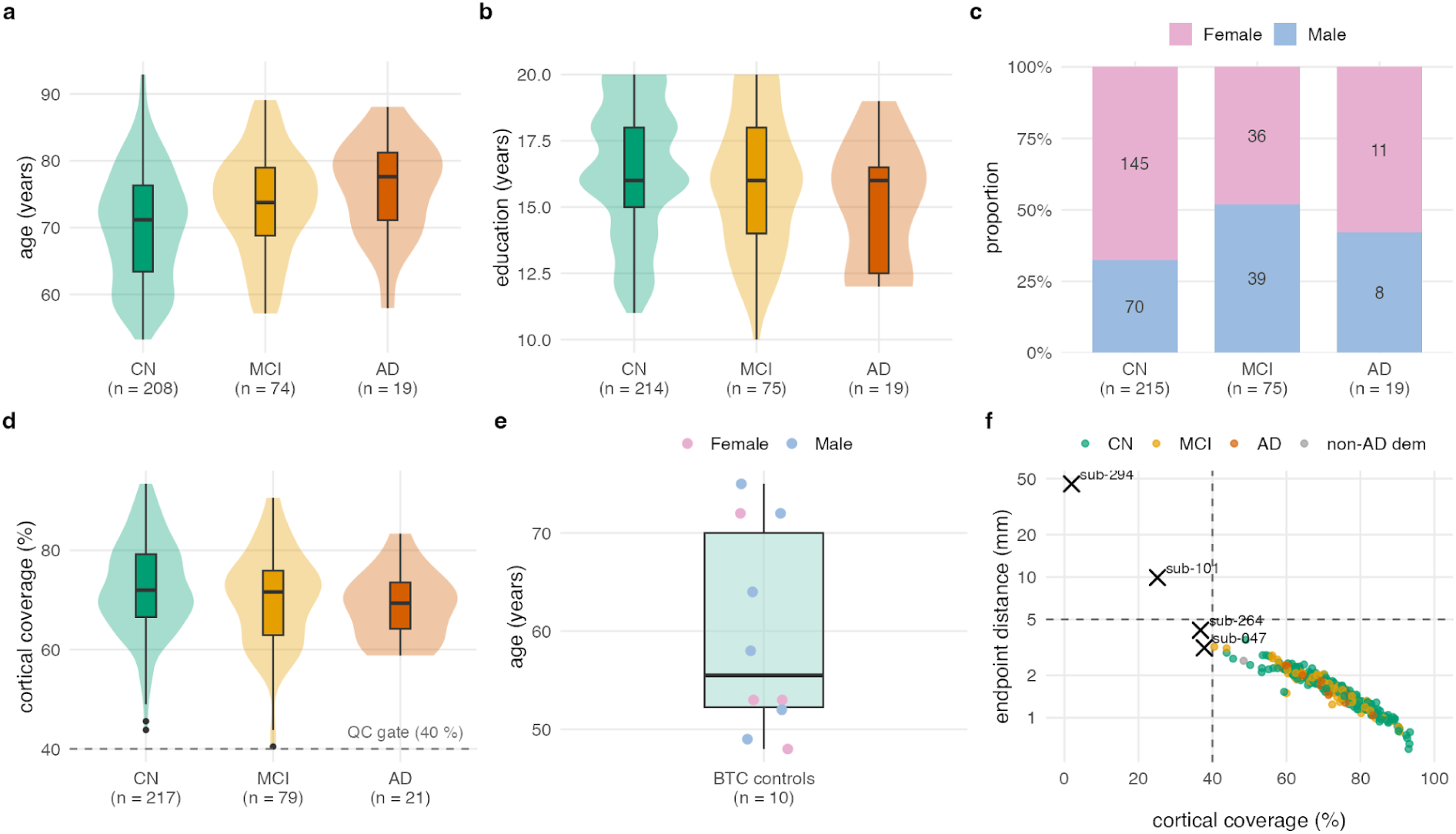
Cohort characteristics and connectome quality-control criteria. (a–c) Demographic characteristics of the ADNI-4 cohort across diagnostic groups: (a) age, (b) years of education, and (c) sex distribution. (d) Cortical coverage, defined as the percentage of white-surface vertices reached by streamline endpoints, in cognitively normal (CN) participants, participants with mild cognitive impairment (MCI), and participants with Alzheimer’s disease (AD) dementia. The dashed horizontal line indicates the 40% cortical-coverage threshold used for quality control. (e) Age and sex distribution of the healthy BTC controls used for method development. Each point represents one participant, colored by sex; the boxplot summarizes age across participants. (f) Relationship between cortical coverage and median streamline endpoint distance from the white surface for the initial ADNI-4 sample, colored by the diagnostic group. Dashed lines indicate the inclusion thresholds of 40% cortical coverage and 5 mm median endpoint distance. Crosses and labels identify participants who failed one or both criteria. Participants with non-AD dementia are shown separately. CN, cognitively normal; MCI, mild cognitive impairment; AD, Alzheimer’s disease dementia; BTC, Brain Tumor Connectomics pre-operative dataset; QC, quality control.

### Subject-specific structure requires multishell acquisition

To determine the acquisition conditions under which subject-specific connectivity can be recovered, we degraded the BTC data of the 10 controls along two axes: streamline count and shell sampling. We measured identifiability, *Idiff*, defined as the similarity between each participant’s degraded and full-data connectivity fingerprints minus the mean similarity to other participants’ fingerprints (Amico and Goñi 2018). Fingerprints were defined as the SIFT2-weighted connectivity of each vertex to vertices within 14.5mm, averaged within 360 parcels for the comparison between participants (Fig. 2).

Streamline count had a modest effect: subsampling the 5-million-streamline tractograms to 100,000 streamlines reduced *Idiff* from .63 (95% CI .60 to .66), the ceiling at which each reference is compared with itself, to .54 (95% CI .51 to .58). All participants remained identifiable from every subsample, and two independent subsamples of the same tractogram identified each other at every count (*Idiff* .46 at 100,000 to .62 at 2 million streamlines).

Shell sampling had a stronger effect. With each degraded acquisition reprocessed from the diffusion signal onward, *Idiff* was .62 (95% CI .59 to .66) with three shells, .50 (95% CI .45 to .54) with two, and .28 (95% CI .19 to .37) with one, and .27 (95% CI .18 to .37) when the angular directions were halved; the three-shell value represents the metric’s ceiling because each full-data fingerprint was compared with itself. Removing each shell reduced *Idiff* in every participant (paired Wilcoxon signed-rank test, *p* = .002 for each removal); *p* = .32 for halving the directions. Single-shell acquisition degraded, but did not erase, individual identifiability: similarity to each participant’s own reference decreased from .86 with two shells to .56 with one, while similarity to other participants decreased from .36 to .28. This loss coincides with the transition at which a single shell no longer supports three-tissue constrained spherical deconvolution, so that tissue compartments can no longer be separated when fiber orientations are estimated. It therefore reflects a joint limitation of the acquisition and the model, rather than the complete loss of individual distinctiveness. The reduction in identifiability was also observed when short-range connectivity was defined using alternative distance thresholds (Supplementary Materials S2).

The ADNI-4 protocol (three shells, b = 500/1000/2000s·mm^-2^, 114 diffusion directions, 2.0mm isotropic) lies on the multishell side of this transition: it matches the BTC reference in shell count and exceeds it in angular sampling and spatial resolution, although its maximum b-value is lower. We therefore applied the method to the ADNI cohort in the analyses that follow.

**Figure 2.**
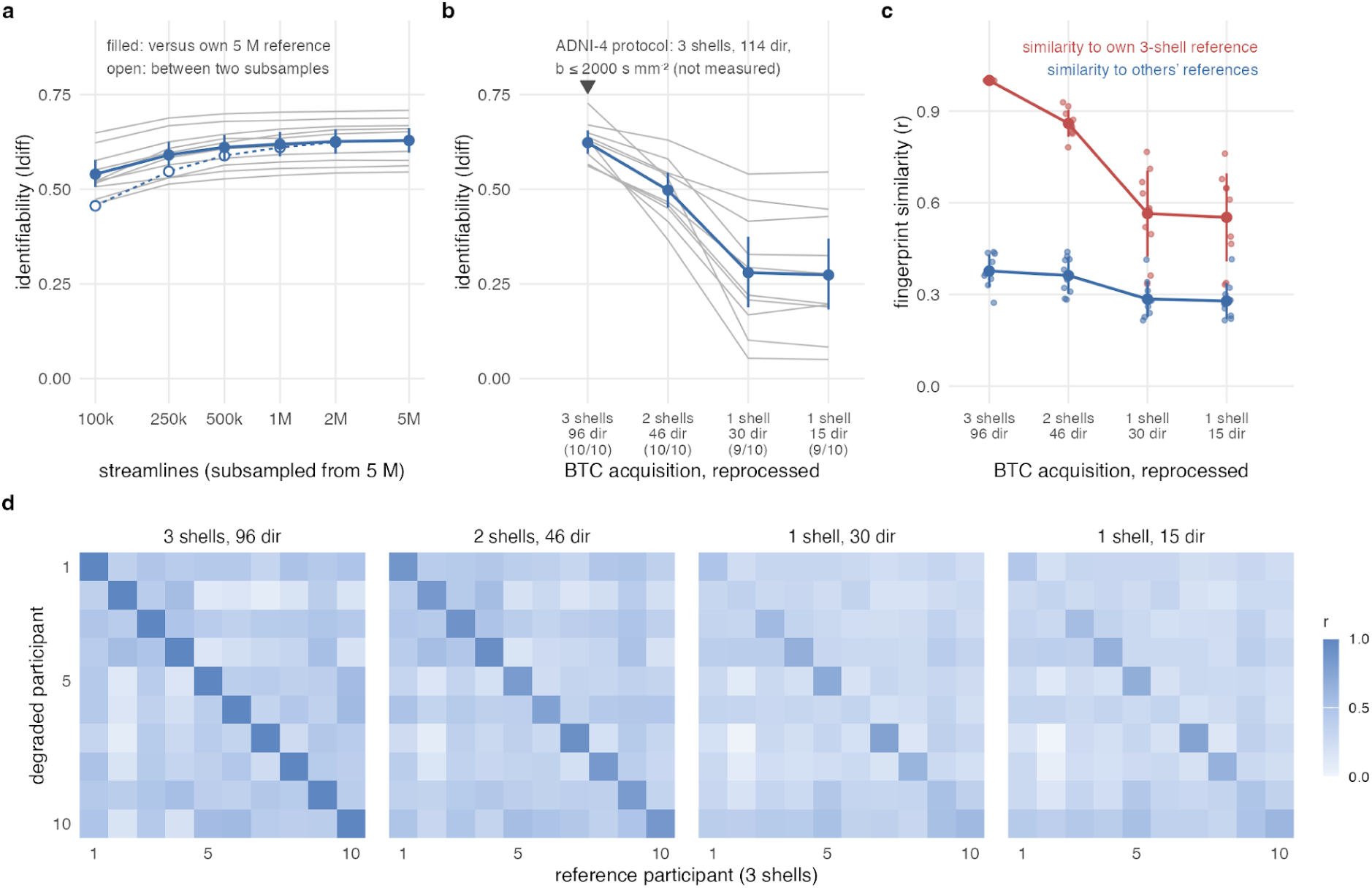
Subject-specific structure is robust to streamline subsampling but degrades with single-shell acquisition. Identifiability of short-range connectivity fingerprints, defined as SIFT2-weighted connectivity between each vertex and vertices within 14.5 mm, averaged within the 360 Glasser parcels. Differential identifiability I_DIFF_ was calculated as the Pearson correlation between each participant’s degraded fingerprint and their full-data reference, minus the mean correlation with other participants’ references. (a) Streamline count. Random subsamples of each 5-million-streamline tractogram (three subsamples per count, averaged). Grey lines, individual participants; filled points, mean I_DIFF_ with 95% confidence intervals from 10,000 participant-level bootstrap resamples; open points, I_DIFF_ between two independent subsamples at the same streamline count. (b) Shell sampling. Each acquisition was reprocessed from response-function estimation through constrained spherical deconvolution, tractography (1 million streamlines), and SIFT2 after removing shells and then halving the remaining directions. Acquisition configurations were three shells (b = 700/1200/2800 s mm^-2^, 96 directions), two shells (b = 700/1200 s mm^-2^, 46 directions), one shell (b = 1200 s mm^-2^, 30 directions), and one shell with half the directions (15 directions). The three-shell condition compares each reference fingerprint with itself and therefore represents the ceiling of I_DIFF_. Numbers in parentheses indicate the number of participants whose degraded fingerprint was most similar to their own reference. Consecutive conditions were compared using paired, two-sided Wilcoxon signed-rank tests (p = .002, .002, and .32). The triangle indicates the ADNI-4 acquisition protocol (three shells, b = 500/1000/2000 s mm^-2^, 114 directions, 2.0-mm isotropic resolution); ADNI-4 data were not degraded. (c) Decomposition of I_DIFF_ into similarity to each participant’s own reference (red) and mean similarity to other participants’ references (blue). Small points, individual participants; large points, mean ± SD. (d) Participant-by-participant similarity matrices for each acquisition condition; rows represent degraded fingerprints and columns represent three-shell reference fingerprints (n = 10).

### A connection-distance hierarchy organizes the vertex-wise structural connectome

Cortical connections span a broad range of lengths, from short links between neighboring regions to long-range tracts, and we first asked how this range is distributed, deriving the band boundaries from the data rather than imposing *a priori* thresholds. In the 217 CN participants who passed registration quality control, we randomly sampled 3 million connections per participant, measured the Euclidean distance between endpoint vertices on the white matter (a lower bound on fiber path length), and weighted each connection by its SIFT2 strength. The weighted length distribution was unimodal, peaking at 11-12mm with a heavy tail beyond 50mm. Its shape was highly consistent across participants: the lower and upper weighted tertiles fell at 14.3 ± .6mm and 27.9 ± 2.1mm (mean ± SD), respectively, and were unrelated to cortical coverage. In contrast, the proportion of connectivity weight in the long-range band varied substantially between individuals (ranging from .18 to .46), providing the basis for the individual-level comparisons in the clinical analyses below.

Partitioning the cohort-average weighted distribution into tertiles produced three bands, short (< 14.5 mm), medium (14.5–27.5 mm), and long (> 27.5 mm), carrying .34, .32, and .34 of the total connectivity weight. A Gaussian mixture analysis of long connection lengths did not identify a preferred number of components, suggesting that the three bands provide a practical partition of the distribution rather than distinct, naturally occurring classes. The boundaries also have an anatomical interpretation. Because endpoint separation underestimates fiber path length, connections in the long band have a path length of at least 27.5 mm, whereas the short band falls within the conventional 3-30mm range attributed to short association (U-)fibers (Movahedian Attar et al. 2020; Shastin et al. 2022). However, the boundaries depend on tractography and connectome-construction settings as well as on anatomy; they should therefore be re-derived for each dataset (Fig. 3; Supplementary Materials S3).

**Figure 3.**
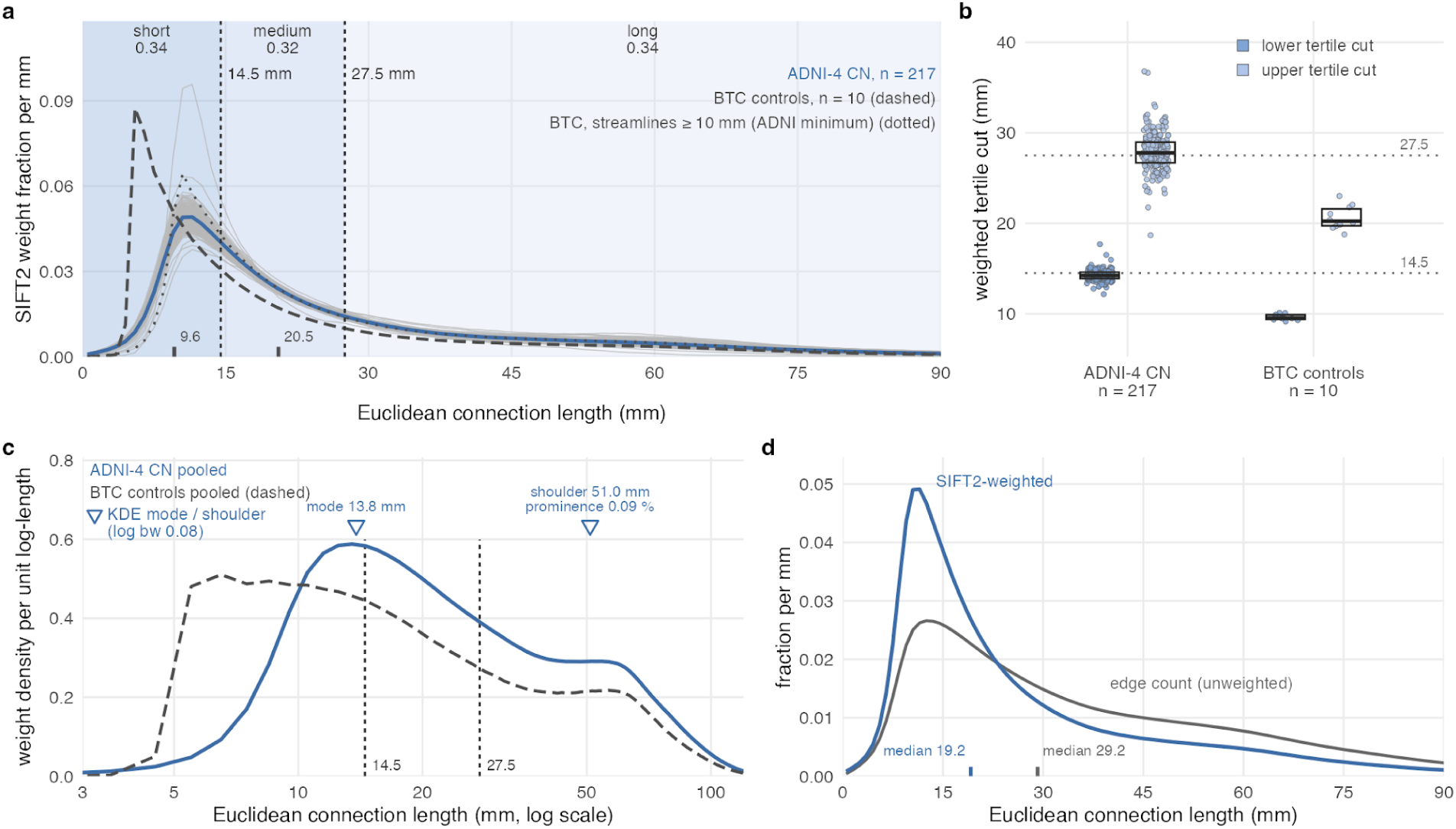
Data-driven partitioning of cortical connections by length. Connection length was defined as the Euclidean distance between endpoint vertices on the white surface in native diffusion space, providing a lower bound on fiber path length. (a) SIFT2-weighted connection-length distributions in 217 cognitively normal ADNI-4 participants who passed registration quality control. Grey lines, individual participants (random samples of 3 million connections per participant, binned at 1 mm and normalized to unit sum); blue line, cohort average. Shading indicates the three bands defined by the weighted tertiles of the cohort-average distribution, with dashed lines at ≈14.5 and ≈27.5 mm marking the corresponding bin centres; values indicate the fraction of total SIFT2 weight in each band. The dashed black line shows the cohort-average distribution for the 10 BTC controls processed using the same procedure, with ticks indicating their weighted-tertile boundaries. The dotted line shows the BTC distribution after excluding streamlines shorter than 10 mm, the minimum streamline length used in ADNI-4 tractography. (b) Lower and upper weighted-tertile boundaries for individual participants in the ADNI-4 and BTC cohorts. Points represent participants; boxes show the median and interquartile range; dotted lines indicate the cohort-level ADNI-4 boundaries shown in a. (c) Cohort-average distributions plotted per unit log-length on a logarithmic axis. The ADNI-4 distribution has a single mode at 13.8 mm (triangle; estimated using kernel density estimation of the resampled distribution) and a shoulder at 50–60 mm; no second mode was detected across the tested bandwidths. (d) Cohort-average ADNI-4 connection-length distributions weighted by SIFT2 strength (blue) or connection count (grey). Ticks indicate the respective medians (19.2 and 29.2 mm). Sample sizes were 217 ADNI-4 and 10 BTC participants.

### Distance and scale are coupled in a nested community hierarchy

Cortical communities can be described at many scales, from a few large modules to many small ones. Whether this organizational scale is coupled to connection distance remains unclear. We decomposed each CN connectome (n = 217) into a multi-resolution community hierarchy by Leiden modularity-based detection applied to the strongest 10% of connections across a resolution sweep (γ = .05–5), yielding a coarse-to-fine cascade (cohort mean 2 to 55 modules). Detection used connectivity weight alone and was blind to distance, allowing us to assess whether a relationship between connection distance and organizational scale emerged from the resulting partition.

The hierarchy was strongly nested (mean containment .93). At the lowest resolution, the partition was the hemispheric split in most participants: two modules were detected in 174 of the 217 participants, and both modules were hemisphere-pure in 140. Each finer level was nested within its predecessor (mean containment .92 to .93 across successive transitions beyond the hemispheric step). As scale became finer, modules became progressively shorter-range and more compact in every participant: within-module connection length fell from 25.0 (95% CI 24.8 to 25.2) to 16.4 (95% CI 16.3 to 16.5)mm, radius of gyration fell from 46.5 to 17.7 mm, and the within-module connectivity fraction from .86 to .34 (sign test *p* < .001 for all three measures). The trends in connection length and compactness were unrelated to cortical coverage (|ρ| ≤ .17 at every level). A distance-blind partition thus yields progressively shorter-range and geometrically more local fine modules, consistent with a progressive decrease in average within-module connection length from larger-scale to finer-scale modules (Fig. 4).

**Figure 4.**
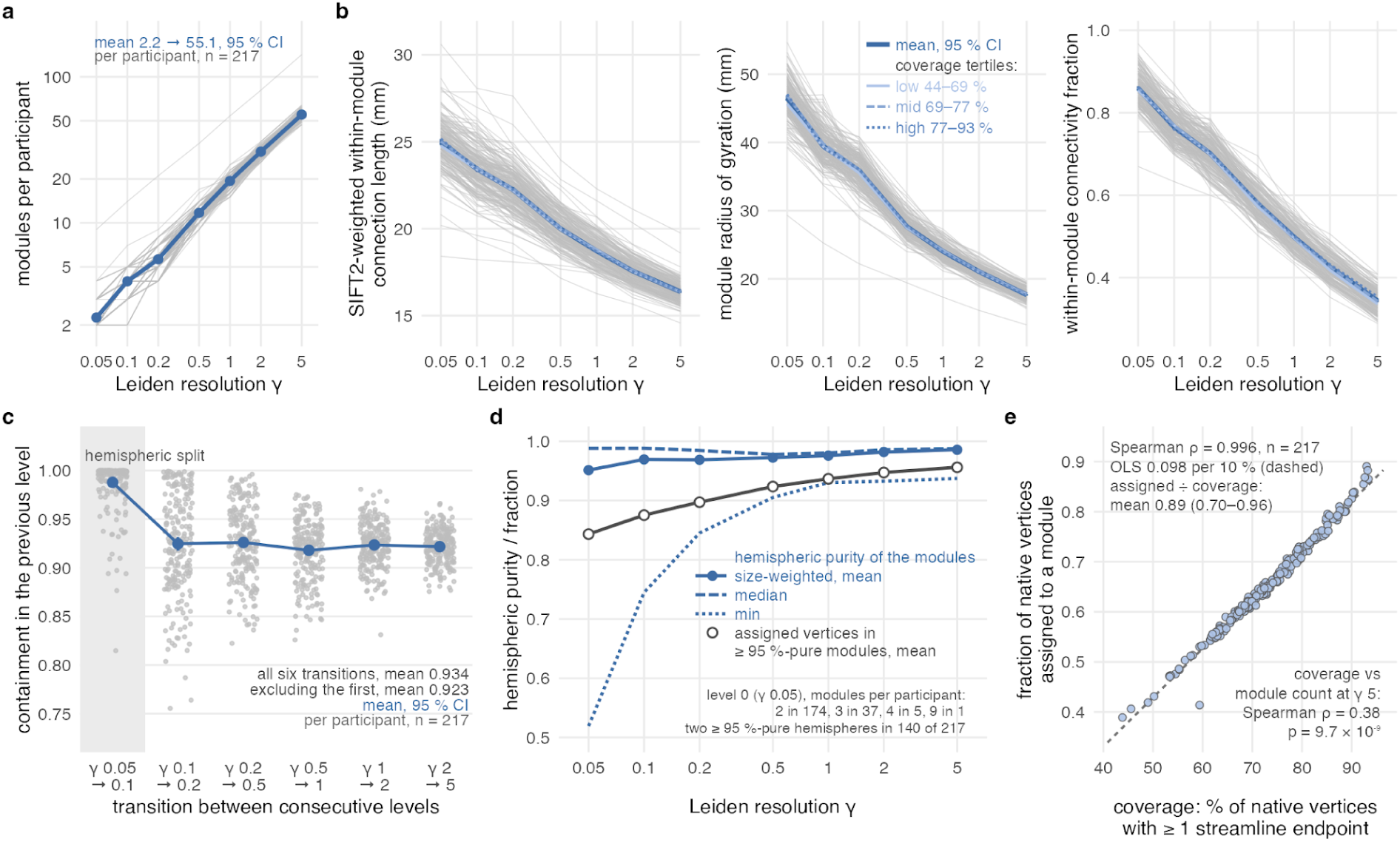
A nested community hierarchy whose modules become shorter-range and more compact with increasing resolution. Leiden modularity detection was applied to the strongest 10% of SIFT2-weighted connections in each vertex-wise connectome at seven resolutions (γ = .05–5) in 217 cognitively normal ADNI-4 participants. (a) Number of modules per participant (grey lines) and cohort mean with 95% confidence interval (CI; blue). (b) SIFT2-weighted within-module connection length (Euclidean endpoint distance), size-weighted module radius of gyration on the native white surface, and within-module connectivity fraction, shown per participant (grey lines) and as cohort means with 95% CIs (blue). In the radius-of-gyration plot, additional lines show means within cortical-coverage tertiles. (c) Containment of each level’s modules within the modules of the preceding level (participants, grey; cohort mean with 95% CI, blue). The first transition, shaded, begins at the lowest resolution, where the partition is the hemispheric split in most participants. (d) Hemispheric purity of modules at each resolution, shown as the size-weighted mean, median and minimum, alongside the fraction of assigned vertices belonging to modules that are at least 95% confined to one hemisphere. Text indicates the distribution of module counts at γ = .05. (e) Fraction of native cortical vertices assigned to a module as a function of cortical coverage, defined as the percentage of native vertices with at least one streamline endpoint. Each point represents one participant; the dashed line shows the ordinary least-squares (OLS) fit. The Spearman correlation and mean OLS-predicted assignment fraction after adjustment for coverage are indicated.

### The hierarchy is an individual fingerprint, strongest in medium- and short-range connections

To establish whether the hierarchy identifies individuals, we compared the reproducibility of each person’s partition across two split halves of their connectome with the similarity of partitions between participants, using the adjusted mutual information (AMI). Split-half reproducibility was high at every level (ranging from .82 to .89), reaching its lowest value at γ = .2 and its highest at the coarsest level, whereas similarity between participants was lower throughout (.57–.66 beyond the coarsest level, which corresponded to the hemispheric split in most participants). The gap, ranging from .18 to .25, was present at every level, with bootstrap 95% CIs excluding zero, and defines an individual fingerprint. Individual identification was achieved in 99.5-100% of participants from γ=.5 onward, and in 75-90% of participants at the three coarse levels. Because the two halves share the same acquisition, registration and cortical surface, split-half reproducibility represents a ceiling for test-retest reliability rather than a direct estimate of it.

Restricting connectivity to each distance band and comparing matched halves of the vertex-wise band-strength maps showed that every participant was identified from every band. In this sense, medium- and short-range connectivity was more individually distinctive than long-range connectivity at every spatial scale., except for the short band at the largest smoothing kernel (kNN 400; .34 versus .36). For example, *Idiff* on Glasser-360 parcel means was .65 for the medium band, .60 for the short band and .47 for the long band, with bootstrap 95% CIs for the between-band differences excluding zero. This pattern reflected greater between-person similarity in long-range strength maps (between-subject r = .53, compared with .35 for the medium band and .40 for the short band). Identifiability was also higher in participants with lower cortical coverage (ρ = -.52 to .-.60); coverage-adjusted values are therefore reported alongside the unadjusted results (Supplementary Materials S4). The cortical reliability of individual differences in band strength was high and near-uniform across the cortex (vertex-wise ICC .97-.99), although it was slightly lower in paralimbic and medial temporal cortex, where endpoint coverage was the lowest. Reliability did not differ between primary sensory and heteromodal association cortex. The individual fingerprint therefore is strongest in medium- and short-range connections (Fig. 5).

Regional atlases summarize connectivity by averaging it within predefined parcels. To see whether this averaging discards individual structure, we compared the vertex-wise band-fraction maps with the same quantities averaged within Desikan–Killiany (68) and Glasser (360) parcels: band composition varied smoothly within individual parcels, so averaging replaced the graded vertex maps with a single regional value and erased the fine, short-range structure shown above to carry identity. Consistently, connectome identifiability rose monotonically with resolution, from *Idiff* = .10 (DK-68; (Desikan et al. 2006) to .19 (Glasser et al. 2016) to ∼.46 at the vertex scale. That identifiability keeps rising to the vertex scale rather than saturating or degrading is not guaranteed: finer resolution reveals more individual structure only if the acquisition can characterize it reliably, so the monotonic increase demonstrates that diffusion data of this quality resolve individual structure to the vertex scale, not merely that finer parcellations create more apparent distinctions.

The same dependence governed cortical routing: the mean number of intermediate nodes between distant anchor regions grew from 3.3 (DK) to 6.6 (Glasser) to 9.7 at the vertex scale, revealing increasingly detailed multi-step routes at higher spatial resolution. These routes were themselves individual, with within-subject routing similarity (.65) far exceeding between-subject (.29), and a group-average connectome reproduced no individual’s routing (.40).

By contrast, the canonical functional precision-imaging partition (Infomap), applied to the structural connectome, was largely predictable from cortical geometry alone, suggesting that the individual signal resides in the routing between conserved endpoints, not in the partition.

**Figure 5.**
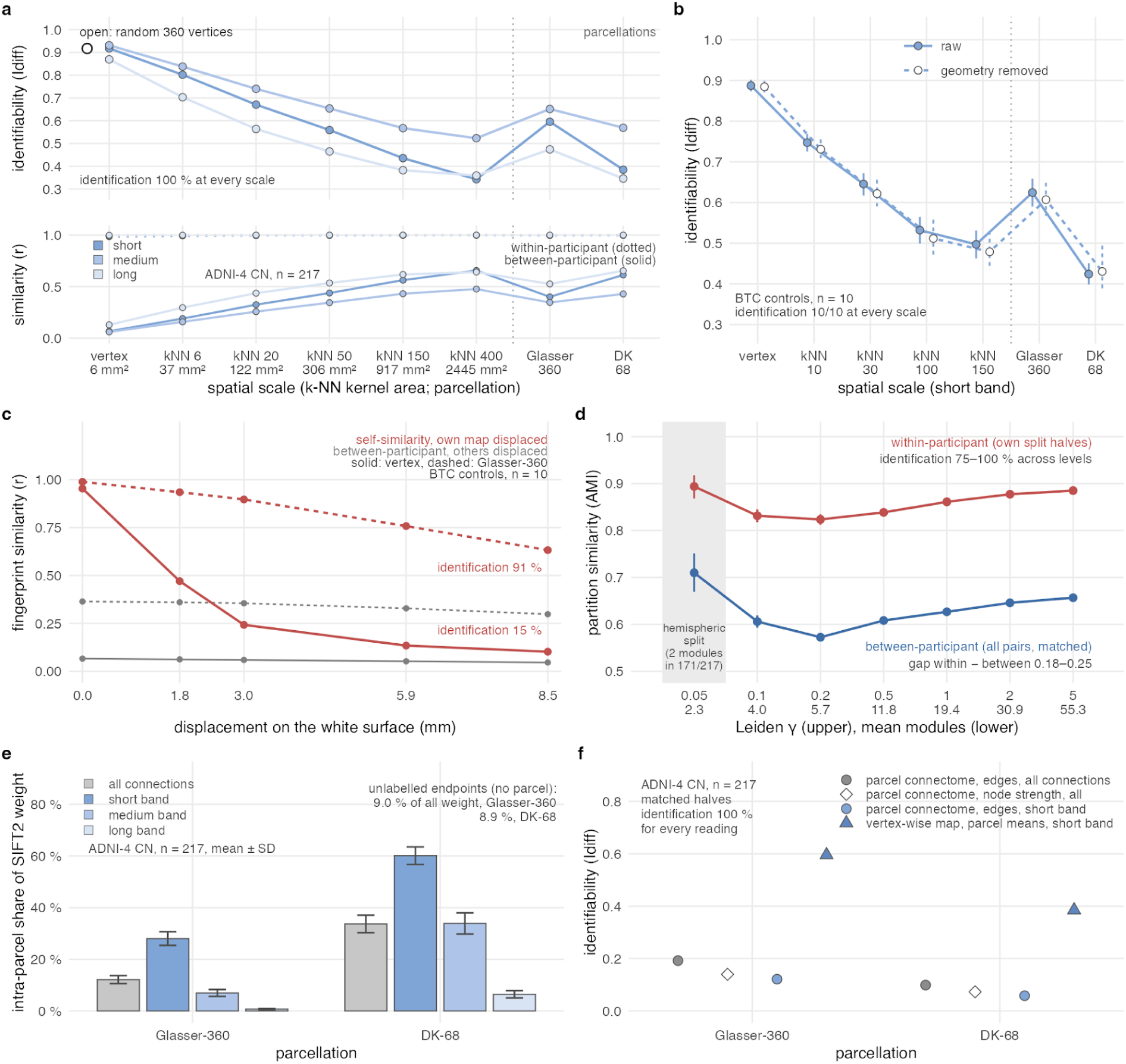
The individual fingerprint across spatial scales and atlases. a) Differential identifiability (I_DIFF_) of vertex-wise band-strength maps in 217 cognitively normal ADNI-4 participants, calculated between matched split halves of each connectome (similarity of half A from one participant to half B from every participant; I_DIFF_ = mean self-similarity minus mean similarity to other participants), for the short, medium and long bands (dark to light). Spatial scales range from unsmoothed fsaverage5 vertices through k-nearest-neighbour (kNN) smoothing kernels (kernel areas shown) to Glasser-360 and Desikan–Killiany-68 parcel means. Points, mean I_DIFF_; error bars, 95% confidence intervals (CIs) from 10,000 participant-level bootstrap resamples, narrower than the points. The open point marks a random subset of 360 vertices for the short band. Every participant was identified at every scale. Lower panel, mean within-participant (dotted) and between-participant (solid) similarity. b) The same analysis for the short-range band in 10 BTC controls, using two independent 2-million-streamline subsamples of each 5-million-streamline tractogram as split halves, before (filled) and after (open, dashed) regressing each participant’s map on their own curvature, sulcal depth, cortical thickness and endpoint-coverage indicator. All 10 controls were identified at every scale. c) Sensitivity to misalignment in the BTC controls. Similarity between a participant’s two halves when one half was displaced by a rigid rotation of the sphere (red; solid, vertex scale; dashed, Glasser-360), and similarity to other participants when their maps were displaced (grey). Values are means over 10 random rotations, drawn independently per hemisphere. The x-axis shows the realised displacement on the fsaverage white surface; percentages indicate identification accuracy at the largest displacement. d) Community-partition fingerprint: adjusted mutual information (AMI) between the two split-half partitions of each participant (red) and between partitions from different participants (blue; all 23,436 unique participant pairs, with both split-half matchings) at each Leiden resolution. Both partitions were projected to fsaverage5, and each split half was thresholded to the same edge density. Points show means; error bars, 95% participant-level bootstrap CIs. Shading marks the lowest-resolution level, which corresponds to a hemispheric split in most participants. e) Share of total SIFT2 weight on connections whose two endpoints fall within the same parcel, for the Glasser-360 and Desikan–Killiany-68 atlases, for all connections and for each distance band (mean ± SD over the 217 participants). Connections with an endpoint outside the atlas labels (approximately 9% of weight for either atlas) are not counted as intra-parcel connections. f) Differential identifiability (I_DIFF_) from the same matched split halves, comparing different representations of the Glasser-360 and Desikan–Killiany-68 atlases: the parcel-by-parcel connectome (edge weights, all connections), its node strength, and the parcel connectome restricted to short-range connections, alongside parcel means of the vertex-derived short-band strength map. Points show mean I_DIFF_; error bars, 95% CIs from 10,000 participant-level bootstrap resamples. Every participant was identified under every representation.

### Individual-referenced short-to-long balance is associated with cognitive and affective variation across the Alzheimer’s continuum

Having established that vertex-wise structural connectivity preserves reliable individual differences, we next examined whether these differences capture clinically meaningful differences in cognitive and affective functioning. Because individuals within the same diagnostic category can differ substantially in cognitive performance and depressive symptoms, we assessed these associations beyond group-average patterns. We therefore referenced every participant against a cognitively normal model rather than against the patient group. We focused on the short-to-long balance, which captures the relative contribution of short- and long-distance connections to each participant’s structural connectome. At each vertex, connectivity was reduced to the short-to-long balance, defined as the relative streamline strength of the shortest and longest distance tertiles (short band < 14.5 mm, within the range of short association U-fibers; long band > 27.5 mm, including longer association and projection pathways), and smoothed over a surface area of approximately 917mm^2^. Each participant’s short-to-long balance was compared with the balance expected from the normative model fitted to the 208 cognitively normal participants. The difference was expressed as a deviation z-score controlling for age, sex, and cortical coverage (Fig. 6).

The resulting deviation maps were highly reproducible and participant-specific. Correlations between independently estimated maps from the same participant were .91 before smoothing and .99 after smoothing. All 301 participants could be identified from either estimate. Despite this strong individual-level reproducibility, the deviations showed little consistent spatial pattern when averaged across the cohort. Nevertheless, they were associated with diagnostic status after accounting for the site. Patients showed higher short-range share than CN participants, with five significant clusters across the precuneus, superior frontal, and lateral temporoparietal cortex (largest *p_FWE_* = .0004; cortex-wide partial *r* = .18, 95% CI .04–.31). The AD-versus-CN contrast also identified three significant clusters (largest *p_FWE_* = .001). Neither contrast was significant without site adjustment.

We next examined whether these individual differences were associated with cognitive performance and depressive symptoms. In pooled analyses, short-to-long balance was associated with episodic memory (six clusters; largest *p_FWE_* = .005), memory reserve (five clusters; largest *p_FWE_* = .008), and CDR-SB (four clusters; largest *p_FWE_* = .005). Higher short-range share was associated with poorer episodic memory and lower memory reserve, as well as greater CDR-SB scores. However, these associations were no longer significant after adjusting for the diagnostic group, and none was significant when analyses were restricted to cognitively normal participants or patients, except for CDR-SB within cognitively normal participants (one cluster, *p_FWE_* = .043). This pattern suggests that the cognitive associations largely reflected differences along the diagnostic continuum rather than associations detectable within diagnostic groups.

The association with depressive symptoms showed a different pattern. Geriatric Depression Scale (GDS) scores were associated with the short-to-long balance across two clusters (largest *p_FWE_* = .0008). Importantly, this association remained significant after adjusting for diagnostic group, with one cluster in the left superior parietal/sensorimotor cortex (*p_FWE_* = .003; Holm-corrected p = .022; partial r = .15, 95% CI .06 to .26). The association was not significant in analyses restricted to either cognitively normal participants or patients, and there was no evidence of an interaction between GDS scores and the diagnostic group (p = .90). This suggests that the association with depressive symptoms was not fully accounted for by diagnostic classification, although it was not demonstrable within either diagnostic subgroup. In contrast, perceived stress showed no significant association (largest *p_FWE_* = .18). Results across all clinical outcomes and analysis groups, together with the sensitivity of diagnostic contrasts to acquisition-site adjustment, are summarized in Supplementary Materials S5.

**Figure 6.**
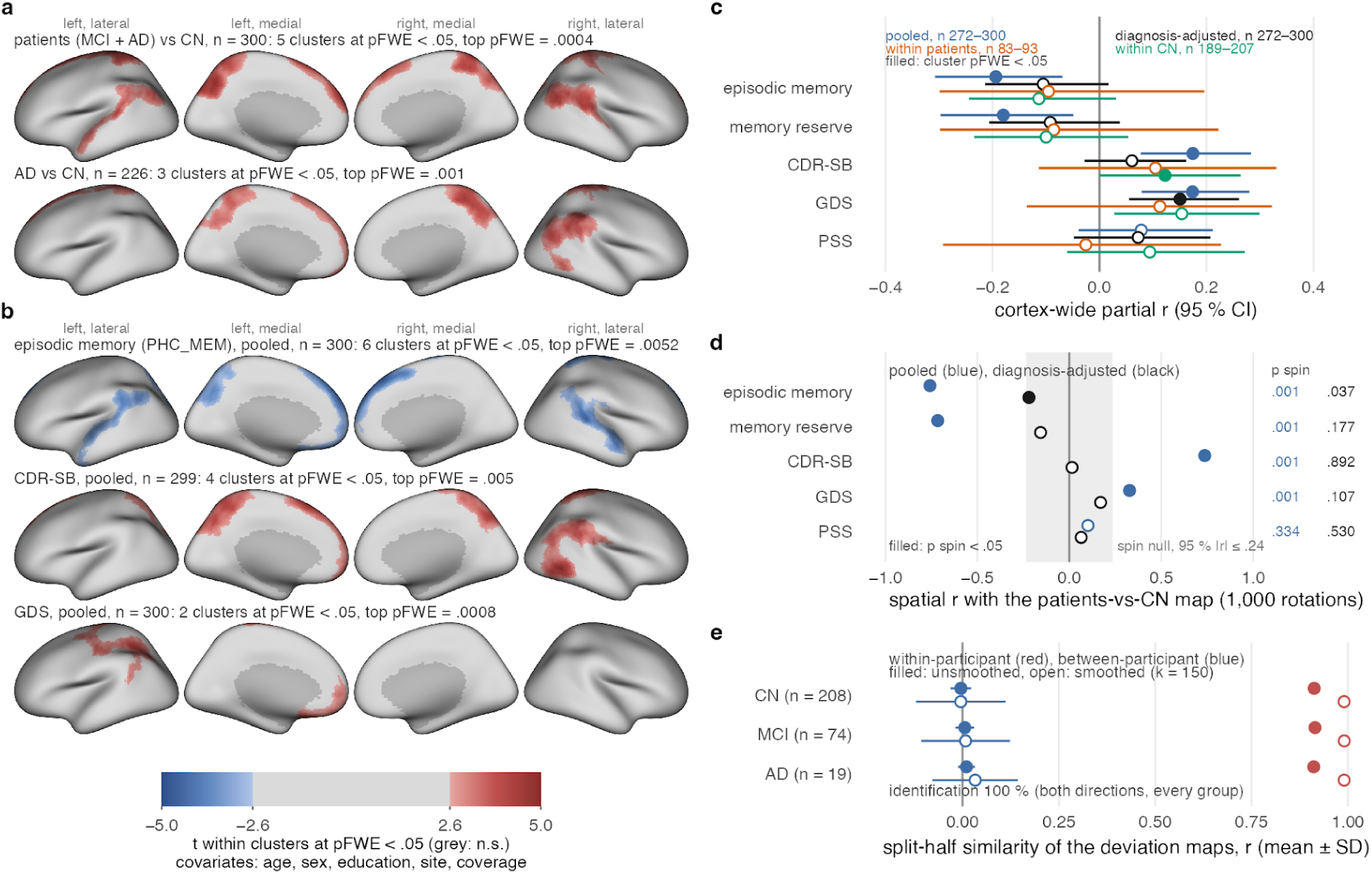
An individually referenced short-to-long balance captures cognitive and affective variation across the Alzheimer’s continuum. a) Matched group contrasts of the short-band deviation z (patients, MCI and AD, versus CN; n = 300; AD versus CN, n = 226), controlling for age, sex, education, site and cortical coverage. t-values are shown within clusters significant at p_FWE_ < .05 (Freedman–Lane cluster-mass permutation, 5,000 permutations, cluster-forming |t| > 2.6); grey indicates regions outside significant clusters. Maps are displayed on the inflated fsaverage surface, with lateral and medial views. b) Pooled vertex-wise models of episodic memory (PHC_MEM), CDR-Sum of Boxes and GDS, using the same covariates and display threshold. c) Cortex-wide partial correlations between the deviation and each outcome (95% CI) from the pooled, diagnosis-adjusted, within-patient and within-CN models. Filled points indicate models with at least one cluster significant at p_FWE_ < .05. d) Spatial correlation of each outcome’s pooled (blue) and diagnosis-adjusted (black) t-map with the patients-versus-CN map; p-values are from spin tests with 1,000 rotations and are shown for p < .05. Shading indicates the central 95% of the spin null distribution. e) Split-half reliability of the deviation maps by diagnostic group: within-participant (red) and between-participant (blue) correlations between maps obtained from the two halves of each connectome, before (filled) and after (open; k = 150) smoothing. Points show mean ± SD.

### The individually referenced balance tracks Alzheimer’s pathology burden beyond diagnosis

Having established that the individually referenced short-to-long balance captures differences in clinical functioning, we next tested whether it also tracked the underlying molecular pathology of Alzheimer’s disease. We related each participant’s vertex-wise short-range deviation to five markers spanning the Alzheimer’s pathological cascade: amyloid burden (PET centiloids), plasma p-tau217, tau-PET, neurofilament light (NfL), and glial fibrillary acidic protein (GFAP). To capture their shared contribution, we also derived a pathology-burden composite from the first principal component of these five markers in the 168 participants with complete data (116 CN, 42 MCI, and 10 AD; 58% of variance explained; loadings .40 to .53). All associations were tested using vertex-wise cluster-mass permutation, controlling for age, sex, education, site, and cortical coverage.

The short-to-long balance was associated with the composite pathology burden across the cohort. Higher pathology burden was associated with a higher short-range share across six clusters, with the largest over the right inferior parietal and superior temporal cortex (largest *p_FWE_* = .001; cortex-wide partial r = .28, 95% CI .11 to .44). Importantly, this association was not explained entirely by diagnostic differences: it remained significant after adjusting for diagnostic group, although it was spatially more restricted, with one cluster over the right inferior parietal and middle temporal cortex (*p_FWE_* = .021; partial r = .21, 95% CI .01 to .38). The association was also detectable within patients (three clusters, *p_FWE_*= .004; n = 52), but not within cognitively normal participants (n = 116; partial r = .06).

The association was not driven by one particular pathology marker. When examined individually in the same participants, the five markers showed similar cortex-wide partial correlations with the short-to-long balance, ranging from .19 to .23. None of the pairwise differences was significant (all Steiger p ≥ .59). Thus, the vertex-wise structural connectome was associated with their shared pathological burden, rather than showing a preferential relationship with any one marker. These results do not support a specific ordering of the associations along the pathological cascade. The strength of this association depended on accounting for site differences. Without site adjustment, the cortex-wide association was negligible (partial r = −.02), whereas adjustment for site alone yielded a positive association (r = .31).

The association was nevertheless consistent across the cohort: it remained significant when each of the 19 sites was excluded in turn, with partial correlations ranging from .22 to .33 and confidence intervals excluding zero. It also remained detectable when site effects were incorporated into the normative model (*p_FWE_* = .041 after adjustment for diagnostic group).

Finally, we assessed whether the time between plasma collection and imaging influenced the result. In 38% of participants contributing to the composite, archived plasma had been collected more than two years before the imaging visit. Restricting the analysis to the 104 participants whose plasma was collected within two years of imaging preserved the direction of the association, although it no longer reached significance (partial r = .20, 95% CI −.04–.42; Fig. 7).

**Figure 7.**
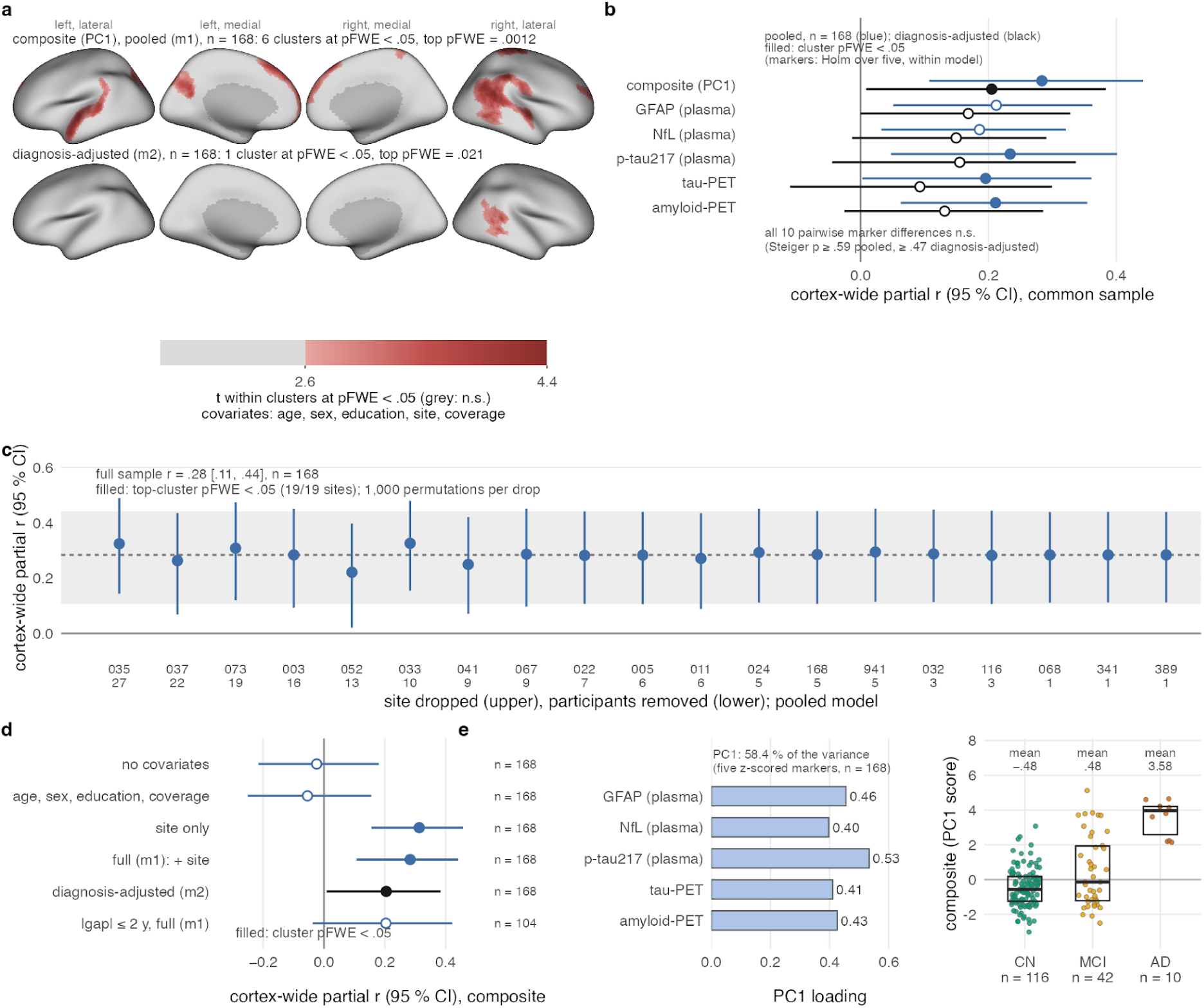
The individually referenced short-to-long balance tracks Alzheimer’s pathology burden beyond diagnosis. a) Vertex-wise association between the short-band deviation z-score and the pathology-burden composite in 168 participants with all five markers, shown for the pooled and diagnosis-adjusted models. Both models control for age, sex, education, site, and cortical coverage. t-values are shown within clusters significant at p_FWE_ < .05 (Freedman–Lane cluster-mass permutation, 5,000 permutations, cluster-forming |t| > 2.6); grey indicates regions outside significant clusters. Maps are displayed on the inflated fsaverage surface, with lateral and medial views. b) Cortex-wide partial correlations (95% CIs from 10,000 bootstrap resamples) between the deviation and the composite and each individual marker, in pooled (blue) and diagnosis-adjusted (black) models. All markers were assessed in the same 168 participants. Filled points indicate at least one cluster significant at p_FWE_ < .05, with Holm correction across the five individual markers within each model. Pairwise comparisons between marker correlations were not significant (Steiger tests). c) Leave-one-site-out analysis of the pooled composite association, excluding each of the 19 sites in turn. The x-axis indicates the site code and number of participants removed; 1,000 permutations were used for each analysis. The dashed line and band indicate the full-sample estimate and its confidence interval; filled points indicate a top cluster significant at p_FWE_ < .05. d) Composite association under different covariate specifications: unadjusted, adjusted for age, sex, education and cortical coverage, adjusted for site alone, fully adjusted pooled model, and diagnosis-adjusted model, alongside the analysis restricted to 104 participants whose plasma was collected within two years of imaging. e) Composite construction, showing the loadings of the five z-scored markers on the first principal component and the resulting component scores across diagnostic groups (boxes show the median and interquartile range; group means are indicated above).

## Discussion

We built a vertex-wise structural connectome to test whether the organization of an individual cortex carries information that group averaging cannot recover. Our results reveal four complementary properties of this representation. First, individual identification depends on the quality of the diffusion data, with multishell acquisition providing the most informative reference. Second, connection distance and community scale are coupled in a nested hierarchy. Third, individual fingerprints are detectable across the spatial scales tested, but their distinctiveness is greatest at the vertex level and is carried preferentially by medium- and short-range connections. Finally, an individually referenced measure of connection-length balance is associated with diagnostic status and Alzheimer’s pathology, while its relationships with cognition, cognitive reserve and depressive symptoms differ in their sensitivity to diagnostic adjustment. Together, these findings establish vertex-wise structural connectomics as a framework for characterizing individual differences in cortical organization. Rather than treating interindividual variation as noise to be averaged away, the framework provides a way to measure that variation and investigate its relationships with clinical and biological characteristics.

Connection distance and community scale are coupled within a nested hierarchy. Coarse, long-range modules form a backbone that nests progressively shorter-range, spatially compact communities, consistent with the multiscale, hierarchically nested community structure described for the connectome (Betzel and Bassett 2017) and with its geometric self-similarity across scales (Zheng et al. 2020). Our contribution is to show that this distance-and-scale fusion holds at the resolution of the cortical surface itself and within single individuals, rather than only in group-level regional graphs. Importantly, this organization was recovered using a partitioning procedure that did not explicitly incorporate connection distance, suggesting that spatial embedding is an important organizing feature of the structural connectome. The hierarchy should consequently be interpreted as a description of the observed organization, rather than evidence that distance and community scale are coupled independently of spatial geometry. This geometric interpretation also provides context for the fingerprinting results: when we applied the functional MRI-derived canonical functional precision-imaging partition (Gordon et al. 2017, 2026; Gratton et al. 2020) to the structural connectome, the resulting modules were largely explained by cortical geometry, suggesting that individual structural information may be expressed not only through differences in community membership but also through the connections linking conserved endpoints.

The individual fingerprint was carried preferentially by short- and medium-range connections and became less distinctive as connectivity was summarized within atlas parcels. This extends structural fingerprinting, previously demonstrated at the voxel (Yeh et al. 2016) and atlas-parcel (Amico and Goñi 2018) levels, and shown to strengthen with increasing resolution (Mansour L et al. 2021), to the cortical surface. Importantly, identification and individual distinctiveness are not interchangeable: participants remained identifiable across the community scales tested, but their connectivity patterns were most distinctive at the vertex level. This loss of distinctiveness with parcellation partly reflects the removal of short-range connections whose endpoints fall within the same parcel. These connections accounted for approximately a third of connectivity weight with the Desikan–Killiany atlas and an eighth with the Glasser atlas. Consistent with this, the same atlas provided a more distinctive fingerprint when applied to vertex-derived maps than when used to construct a parcel-level connectome. Thus, vertex-wise construction preserves individual information that can subsequently be summarized at coarser scales, whereas direct parcel-level construction discards some of that information from the outset.

This finer representation, however, comes with greater sensitivity to anatomical alignment. A displacement of just two millimetres substantially degraded the unsmoothed vertex fingerprint, highlighting how small registration errors can affect individual-level comparisons. This is the structural counterpart of the misalignment problem addressed by hyperalignment in functional neuroimaging (Haxby et al. 2011, 2020; Feilong et al. 2018), and connectivity-guided surface registration (Zhou et al. 2026) may push the usable scale finer. The vertex-wise connectome is best used as the measurement, and comparison between people is best made at kernel areas of a few hundred square millimetres or on parcel means of the vertex-derived maps. The fingerprint also survived the removal of each participant’s cortical geometry, supporting the interpretation that it reflects individual connectivity patterns rather than simply differences in cortical folding.

The clinical analyses make the consequences of individual referencing explicit. Although the individually referenced short-to-long balance provides a reliable characterization of individual connectivity organization, its clinical associations require a qualified interpretation. Matched group contrasts using the same inference framework distinguished participants with Alzheimer’s disease from cognitively normal participants after accounting for acquisition site. Associations with episodic memory, dementia severity and the memory-reserve index were spatially similar to the diagnostic contrast, but none remained significant after adjustment for diagnostic group or was detectable within either subgroup. These associations therefore appear to reflect differences related to diagnostic status rather than variation among individuals within the same diagnostic group. The association with depressive symptoms showed a different pattern: it remained detectable after adjustment for diagnosis, although it was not significant within either subgroup and did not differ between groups. Thus, this association was not fully accounted for by diagnostic classification, although it does not establish a depression-specific effect within either group. Perceived stress, in contrast, was not associated with the measure. The dependence of these clinical effects on acquisition-site adjustment is also important. Diagnostic composition differed across ADNI sites, and site effects were sufficiently strong that no clinical associations were detectable without accounting for site. Acquisition sites should therefore be considered when applying normative models to multisite clinical datasets.

The pathology analysis extends the same logic from behaviour to biology. A composite of amyloid, p-tau217, tau-PET, neurofilament light and GFAP was associated with the individually referenced connectivity measure across the Alzheimer’s continuum, and this association remained detectable after adjustment for the diagnostic group. The association was detectable within the patient group, but not within cognitively normal participants, and was robust to leaving out individual acquisition sites. These findings suggest that individual variation in connection-length organization is related to shared variation across several markers of Alzheimer’s pathology, beyond differences between diagnostic groups. The five markers contributed similarly to the composite, so we cannot establish whether the connectome is preferentially sensitive to any particular component of the biomarker cascade or infer a temporal sequence. The association was weaker in the subset with plasma samples collected within two years of imaging, although uncertainty in the timing of sample collection limits the interpretation of this sensitivity analysis. Accordingly, we cannot conclude that the measure preferentially reflects downstream, structure-damaging pathology. Rather, it appears to capture individual connectivity variation associated with a broader burden of Alzheimer’s pathology.

A practical implication follows from the degradation analysis. The acquisition experiments show that the quality of the individual fingerprint depends on the diffusion information available, with the transition from multishell to single-shell data producing a substantial reduction in similarity to the three-shell reference. Nevertheless, individual identification remained possible under single-shell acquisition, indicating that reduced diffusion information degrades individual-level characterization rather than necessarily eliminating identifiability. ADNI-4 itself uses a two-tier acquisition scheme, reserving the multishell Advanced protocol for multiband-capable scanners and applying a single-shell Basic protocol elsewhere. Consequently, only a subset of ADNI-4 participants acquired with the Advanced protocol supports the multishell analyses presented here, illustrating how acquisition choices can constrain the application of individual-level structural connectomics even within large imaging studies.

The position of ADNI-4 on our acquisition ladder is inferred from its protocol rather than directly measured in this cohort. These findings therefore do not establish a universal minimum acquisition protocol, but quantify the effects of reducing diffusion information under the acquisition and processing conditions tested here. Where individual-level inference is the goal, multishell sampling should be considered at the protocol-design stage, while future methods that recover tissue separability from sparser sampling could extend the approach to a wider range of datasets. Tractography settings also matter for the distance bands. The minimum streamline length alone shifted the reference cohort’s tertile cut-offs from 9.5 and 20 mm to the values used in ADNI, indicating that distance thresholds should be re-derived for each dataset and processing pipeline rather than assumed to transfer directly between studies.

Several limitations should be considered when interpreting these findings. First, vertex-wise connectomes require accurate cortical surface reconstruction and interindividual alignment, as small correspondence errors can disproportionately affect high-resolution connectivity representations. Second, the connectome remains an indirect representation of white-matter anatomy: streamline endpoints and Euclidean distances approximate cortical connectivity and connection length, while tractography is affected by gyral bias and incomplete cortical coverage. The distance bands should therefore be understood as operational categories rather than direct anatomical classifications, and may require re-estimation for different acquisition and processing pipelines. Finally, the clinical analyses are cross-sectional and rely on a limited set of cognitive and affective measures, restricting conclusions about temporal relationships and the clinical meaning of the observed associations. The framework was also evaluated using high-quality diffusion data, and its performance across datasets with different acquisition protocols and image quality remains to be established.

In conclusion, vertex-wise structural connectomics preserves individual differences in cortical organization that are lost through group averaging and atlas parcellation. Our findings demonstrate its potential for characterizing individual connectivity patterns across spatial scales and relating them to clinical and biological variation, while highlighting the importance of diffusion-data quality and accurate anatomical correspondence. By treating interindividual variability as a signal rather than noise, this framework opens new opportunities to investigate how individual differences in structural brain organization relate to cognition, affect and health.

## Methods

### Study design and cohorts

We aim to recover the structure that distinguishes one brain from another and to relate it to a person’s clinical and affective state. Standard connectomics averages connectivity into atlas parcels and compares group means, which removes the very individually-shifted features that carry this information. We therefore adapt the principle of precision imaging, established for functional connectivity (Gordon et al. 2026, 2017), to the structural connectome: every cortical surface vertex is treated as a node, and each brain is characterized on its own terms rather than pooled. The study uses two independent cohorts under a frozen-template design, so that the method is fixed before it is applied and the application cannot be tuned to its result. On the reference cohort, the BTC (Aerts et al. 2022), a high-quality multi-shell acquisition, we develop the method and establish the data quality it requires: by progressively degrading these acquisitions and reprocessing each end to end, we determine the minimum quality at which the subject-specific measures remain reliable, which lets us decide in advance whether a given clinical dataset qualifies. We then apply the fixed method to the clinical cohort, ADNI-4 (Jack et al. 2008), which meets this requirement, to recover subject-specific structure across the Alzheimer’s continuum. To make this structure individual rather than relative to the patient group, we reference each brain against a normative model built from cognitively-normal participants: at every vertex we express connectivity as the balance between short-range (U-fiber-range) and long-range connections and score how far a person departs from the value expected for their age and sex, so that individual deviations from this healthy reference, not differences between group means, become the unit of analysis.

### Datasets

Data used in the preparation of this article were obtained from the Alzheimer’s Disease Neuroimaging Initiative (ADNI) database (adni.loni.usc.edu). The ADNI was launched in 2003 as a public-private partnership, led by Principal Investigator Michael W. Weiner, MD. The original goal of ADNI was to test whether serial magnetic resonance imaging (MRI), positron emission tomography (PET), other biological markers, and clinical and neuropsychological assessment can be combined to measure the progression of mild cognitive impairment (MCI) and early Alzheimer’s disease (AD). The current goals include validating biomarkers for clinical trials, improving the generalizability of ADNI data by increasing diversity in the participant cohort, and providing data concerning the diagnosis and progression of Alzheimer’s disease to the scientific community. For up-to-date information, see adni.loni.usc.edu. The present analyses used data from the fourth phase of the study (ADNI-4), downloaded in February 2026.

The Brain Tumor Connectomics pre-operative dataset (BTC) is an openly available multimodal 3 T MRI dataset on OpenNeuro that pairs high-quality, multi-shell diffusion imaging with structural and functional acquisitions in pre-operative brain-tumour patients and healthy control subjects (Aerts et al. 2022, 2018). The group structural template, the connectivity-gradient alignment basis, and the reliability (acquisition-degradation) analyses were derived from the 10 healthy controls. The clinical cohort was drawn from the ADNI, a multi-site study of older adults that spans cognitively normal, mild cognitive impairment, and AD.

### Image acquisition and preprocessing

For both cohorts, the raw inputs were T1-weighted anatomical and diffusion-weighted (DWI) acquisitions; detailed information about both MRI data acquisition sequences can be found in the supplementary materials S6. The T1-weighted image was processed with FreeSurfer 8.2.0 (Fischl 2012) to reconstruct the cortical surfaces, which served as the structural reference onto which connectivity was later projected. The DWI data was preprocessed using the MRtrix3 (Tournier et al. 2019) toolbox. Fiber orientation distributions were estimated by multi-shell multi-tissue constrained spherical deconvolution (MSMT-CSD; (Jeurissen et al. 2014), and anatomically-constrained tractography (ACT; (Smith et al. 2012) was performed with second-order integration over fiber orientation distributions (iFOD2). The five-tissue-type segmentation required by ACT was generated from the FreeSurfer 8.2.0 reconstruction.

For each subject, a whole-brain tractogram was generated and weighted by SIFT2 (Smith et al. 2015) In ADNI-4, 10 million streamlines were generated using dynamic seeding, with backtracking and cropping at the grey–white matter interface, and a minimum streamline length of 10 mm. In BTC, 5 million streamlines were generated by seeding from the grey–white matter interface, with a minimum streamline length of 5 mm. Both tractograms were weighted using SIFT2, allowing streamline contributions to be quantitatively weighted when constructing the connectome.

### Vertex-wise structural connectome

For each subject, a vertex-wise structural connectome was built in three stages: (i) registration, (ii) surface projection, and (iii) endpoint-to-vertex aggregation.

First, for each participant, the T1-weighted volume was registered to the subject’s DWI space using a hybrid procedure. A boundary-based registration (BBR) was computed using FSL (Jenkinson et al. 2012), and mutual-information FLIRT registration was computed as an independent candidate. Then, we selected the transform that minimized the median distance between streamline endpoints and the cortical surface, thereby choosing the registration that best aligned tractography with anatomy on a per-subject basis.

Second, the FreeSurfer white-matter surfaces of both hemispheres were projected into DWI space using the selected transform. Vertex-wise parcel labels were taken from the surface from the Desikan–Killiany atlas (Desikan et al. 2006), which is registration-independent. The resulting surfaces comprised approximately 244k to 291k vertices per subject. The vertex count varies across subjects because these are each subject’s native FreeSurfer tessellation rather than a resampled common mesh; cross-subject correspondence is not assumed at this stage but is established downstream by projecting each subject onto the shared fsaverage template in the next steps.

Third, each streamline endpoint was assigned to cortical vertices using the Mahalanobis multi-vertex scheme. Each endpoint contributed to its top-k = 5 Mahalanobis-nearest vertices within a search radius of two standard deviations (σ_n_ = σ_t_ = 2mm), with a contribution weighted by 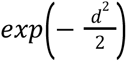. These weights define two sparse endpoint-assignment matrices 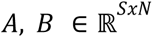, where *S* is the number of streamlines, *N* the number of cortical vertices, and *A_su_* (respectively *B_sv_*) represent the Mahalanobis assignment weight linking the first (respectively second) endpoint of streamline *s* to vertex *u* (respectively *v*), with at most k non-zero entries per row. Let *W* = *diag*(*w*) be the diagonal SIFT2 streamline weights *w*_s_. The symmetric vertex-by-vertex connectivity matrix is then the product of:

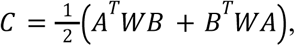

So that each edge *C*_uv_ accumulates the SIFT2-weighted streamline density assigned between vertices *u* and *v*.

Furthermore, for each subject we also create an anatomy file containing vertex-wise cortical thickness, surface area, and fractional anisotropy (FA), the Desikan-Killiany labels, and the registered spherical and inflated surface geometries, so that all downstream analyses could run without further FreeSurfer dependencies.

### Network Hierarchy

The connection-distance bands were derived from the cognitively normal cohort. For each participant, we sampled 3 million connectome edges with replacement and computed the Euclidean distance between their endpoints on the native white-matter surface, using this distance as a lower bound on fibre length (Roberts et al. 2016; Betzel et al. 2016). Each distance was weighted by the corresponding SIFT2 streamline strength. The resulting weighted distance distributions were binned at 1-mm intervals, normalized to unit sum for each participant, and averaged across participants with equal weight. The cohort-average distribution was then partitioned into weighted tertiles to define short-, medium-, and long-range connection bands.

To characterize the scale of cortical organization, each subject’s connectome was thresholded to its top 10 % of edges by weight and partitioned with the Leiden algorithm under a modularity objective (igraph implementation; (Traag et al. 2019) across a sweep of resolution parameters (γ = .05, .1, .2, .5, 1, 2 and 5; two optimization iterations; modules smaller than 100 vertices removed). Ordering the resulting partitions from coarse to fine produced a nested cascade, the nesting of which was quantified as containment, the mean over finer modules of the largest fraction of a module falling within a single coarser module.

Each level was then summarized with SIFT2-weighted measures computed over a sample of (10^6^) edges: the mean within-module connection length, the module spatial compactness (the size-weighted mean radius of gyration of each module’s vertex coordinates on the inflated surface), the segregation (the fraction of total connectivity contained within modules), and the endpoint-mean fractional anisotropy and cortical thickness along within-module connections. We also quantified the fraction of sampled connections falling within modules. A subject was flagged as a metric outlier when its mean within-module length had an absolute cohort-level z-score greater than 3.5.

### Subject-specific measures

Before applying the method to ADNI, we established the acquisition requirements for recovering subject-specific structure by degrading the high-quality BTC acquisitions from 10 healthy controls along two axes and reprocessing each degraded dataset end-to-end, from response-function estimation through constrained spherical deconvolution, tractography and SIFT2. One axis reduced the streamline count, using random subsamples ranging from 100,000 to 2 million streamlines from each 5-million-streamline tractogram (three random subsamples per condition). The other reduced diffusion quality by dropping shells and angular directions, from the full multishell acquisition to two shells or a single shell with either 30 or 15 directions. For each degraded dataset, we computed differential identifiability, defined as the difference between the mean similarity of each participant’s degraded fingerprint to their own full-data reference and the mean similarity to the full-data references of other participants. Fingerprints were based on short-range (< 14.5 mm) connectivity strength aggregated to the Glasser-360 parcellation (Glasser et al., 2016). We also assessed identification accuracy and estimated 95% confidence intervals using 10,000 participant-level bootstrap samples.

The reliability of the hierarchy within a subject was measured by splitting each connectome into two disjoint edge sets, partitioning each independently across the resolution sweep, and computing the adjusted mutual information (AMI) between the two partitions at every level. Between-subject similarity was measured by computing the AMI between partitions from different subjects after projection to the fsaverage5 surface. All 23,436 unique between-subject pairs were evaluated in both comparison orders. Identifiability was quantified as the difference between within- and between-subject AMI, and identification accuracy was assessed by determining whether each partition was most similar to the other partition from the same subject.

A vertex-wise individual-dominance map, defined as one minus group typicality, was computed at γ = .5 to localise where subjects diverged most from the group. We further quantified band-restricted identifiability by building a vertex-wise connectivity-strength map for each band, splitting the edges into two disjoint halves, and projecting each half to fsaverage5. Fingerprints were compared at multiple spatial scales: at the native vertex level, after k-nearest-neighbour smoothing (k = 6, 20, 50, 150 and 400), and after aggregation to the Glasser-360 and Desikan–Killiany parcellations. For each scale, identifiability was calculated as the difference between within-subject and between-subject fingerprint correlations. The analyses were also repeated after regressing cortical coverage from each feature.

Vertex-level reliability was additionally assessed using the intraclass correlation coefficient (ICC(1)) between the two split-half connectivity-strength maps across participants. The spatial distribution of reliability was compared across Mesulam cortical classes using spin tests (1,000 rotations per hemisphere; (Alexander-Bloch et al. 2018).

Individual routing was assessed by tracing shortest paths between predefined anchor parcels on each subject’s connectome and counting the intermediate nodes on each route at three spatial resolutions (Desikan–Killiany, Glasser-360 and vertex); routing signatures were then compared within subjects, between subjects, and against a group-average connectome. To test whether the standard precision-imaging partition reflects individual connectivity or cortical geometry, we compared each subject’s Infomap consensus parcellation (Rosvall and Bergstrom 2008) with a geometry-only k-means parcellation of the cortical surface coordinates with the same number of modules, quantifying their agreement by AMI together with the spatial compactness of the Infomap modules.

For the clinical analyses, connectivity at each vertex was expressed as the fraction of its total SIFT2 strength carried by each band, smoothed on the fsaverage5 surface by k-nearest-neighbour averaging (k = 150). A cognitively normal normative model, in which the short-band fraction was modeled as a function of cortical coverage, age and sex, was fitted at each vertex using cognitively normal participants. Each participant’s departure from the expected value was expressed as a deviation z-score, with leave-one-out scoring used for the cognitively normal participants. This individually referenced, fraction-based deviation was the measure used throughout the clinical analyses. The log-ratio of short- to long-range connectivity was examined as a sensitivity measure.

### Group-specific measures

Group-level organization was assessed in two ways. To test whether the disease produces a shared spatial pattern, we computed the signed group-mean deviation map and the case-control contrasts (AD versus cognitively normal, and all patients versus cognitively normal), together with a vertex-wise cancellation index defined as the ratio of the signed group mean to the mean absolute deviation, where zero indicates a purely individual and one a fully shared pattern. To relate the individual deviations to phenotype, we fitted vertex-wise models across subjects testing the association between each participant’s deviation and episodic memory, a memory-reserve index, dementia severity (CDR-Sum of Boxes), depression (Geriatric Depression Scale), and perceived stress (PSS-10). These associations were examined using pooled models, models additionally adjusting for diagnostic group, and separate models within the patient and cognitively normal groups. For depression, we additionally tested the interaction between GDS scores and diagnostic group. The reciprocity of the redistribution was assessed as the spatial correlation between the short- and long-band association maps, with the medium band as a null comparator.

### Alzheimer’s pathology

Alzheimer’s pathology was indexed by five fluid and imaging biomarkers spanning the disease cascade: amyloid burden (PET, centiloids), plasma p-tau217, tau-PET (meta-temporal ROI), plasma neurofilament light (NfL), and plasma glial fibrillary acidic protein (GFAP). Plasma p-tau217, NfL, and GFAP were log-transformed; amyloid and tau-PET were used as provided. To index overall pathology burden, the five markers were standardized across participants with complete data and reduced to their first principal component, oriented so that higher scores indicate greater pathology (n = 116); its variance explained and marker loadings were recorded. This composite was used as the primary measure of Alzheimer’s pathology, with the five individual biomarkers examined in secondary analyses.

At every vertex, we related the individually referenced short-range deviation to the pathology-burden composite and to each individual biomarker. Each association was estimated as a partial correlation after adjusting for age, sex, education, and acquisition site, and tested by the same cluster-mass permutation inference used throughout (cluster-forming (*|t|* > 2.6), 5,000 Freedman–Lane permutations, significance at (*p_FWE_* < .05). Associations were examined in the pooled sample, after additionally adjusting for the diagnostic group, and separately within participants with cognitive impairment and cognitively normal participants.

### Statistical analyses

Vertex-wise association models included age, sex, education and acquisition site (19 ADNI sites as fixed effects) as covariates. The memory-reserve index was calculated as the residual of episodic memory after regression on age, sex, and cortical thickness, an atrophy proxy, so that it captured cognition retained beyond what structural atrophy would predict. Because surface smoothing makes parametric degrees of freedom unreliable, inference used cluster-mass permutation with a max-statistic family-wise correction: a cluster-forming threshold of (t ≈ 2.6) (two-sided (p < .01)), 5,000 permutations to construct the null distribution, and significance at cluster-level (*p_FWE_* < .05). The behavioral orthogonality of affect to the cognition-reserve axis was assessed by partial correlation controlling for the relevant covariates. Community detection and graph measures used igraph and scikit-learn in Python, and surface handling used the FreeSurfer fsaverage template with the Glasser-360 and Desikan–Killiany atlases.

### Toolbox, code and data availability

All code and data used in this study is openly accessible. The DWI preprocessing and tractography building code is available in https://github.com/josepinedamartinez/adni4_vwisecon_dwi, and the vertex-wise pipeline is documented and reproduced in an interactive toolbox at https://saulpascualdiaz.github.io/understanding_vertexwise_connectome. The BTC dataset can be retrieved from OpenNeuro (Aerts et al. 2018). ADNI data is available on the ADNI data portal (https://adni.loni.usc.edu) via application.

## Data Availability

ADNI data are available to qualified researchers through the ADNI data portal (https://adni.loni.usc.edu) upon application and acceptance of the ADNI Data Use Agreement. The BTC dataset is openly available on OpenNeuro (ds001226). DWI preprocessing and tractography code is available at https://github.com/josepinedamartinez/adni4_vwisecon_dwi, the vertex-wise pipeline is documented in an interactive toolbox at https://saulpascualdiaz.github.io/understanding_vertexwise_connectome, and code and derived data to reproduce the figures are available at https://github.com/saulpascualdiaz/publications/tree/main/pineda_2026_vertexwise_connectomics.

## Acknowledgements

Data collection and sharing for the Alzheimer’s Disease Neuroimaging Initiative (ADNI) is funded by the National Institute on Aging (National Institutes of Health Grant U19AG024904). The grantee organization is the Northern California Institute for Research and Education. In the past, ADNI has also received funding from the National Institute of Biomedical Imaging and Bioengineering, the Canadian Institutes of Health Research, and private sector contributions through the Foundation for the National Institutes of Health (FNIH) including generous contributions from the following: AbbVie, Alzheimer’s Association; Alzheimer’s Drug Discovery Foundation; Araclon Biotech; BioClinica, Inc.; Biogen; Bristol-Myers Squibb Company; CereSpir, Inc.; Cogstate; Eisai Inc.; Elan Pharmaceuticals, Inc.; Eli Lilly and Company; EuroImmun; F. Hoffmann-La Roche Ltd and its affiliated company Genentech, Inc.; Fujirebio; GE Healthcare; IXICO Ltd.; Janssen Alzheimer Immunotherapy Research & Development, LLC.; Johnson & Johnson Pharmaceutical Research & Development LLC.; Lumosity; Lundbeck; Merck & Co., Inc.; Meso Scale Diagnostics, LLC.; NeuroRx Research; Neurotrack Technologies; Novartis Pharmaceuticals Corporation; Pfizer Inc.; Piramal Imaging; Servier; Takeda Pharmaceutical Company; and Transition Therapeutics. The BTC_preop dataset is openly available on OpenNeuro (ds001226); we thank its contributors.

## Author contributions

J.P. and S.P.-D. conceived the study and designed the analytic framework. S.P.-D. developed the vertex-wise structural connectome pipeline and the accompanying open toolbox, implemented the normative modeling and permutation-inference code, performed the acquisition-degradation analyses on the reference cohort and supervised the work; J.P. performed the MRI preprocessing, tractography and connectome construction for the clinical cohort; S.P.-D and J.P. carried out the distance-band, community-hierarchy, identifiability, routing, normative-deviation and biomarker analyses, and produced the figures; M.C.-T. contributed to the design, method validation and interpretation of the cognitive reserve and clinical analyses. J.P. and S.P.-D. wrote the first draft of the manuscript. E.S. Proofread the manuscript. All authors revised the manuscript critically and approved the final version.

