## Supplementary materials for "Diffusion-based vertex-wise connectomics encodes individual differences in cognition, affect and pathology"

#### S1. Participant inclusion and sample sizes across datasets and analyses

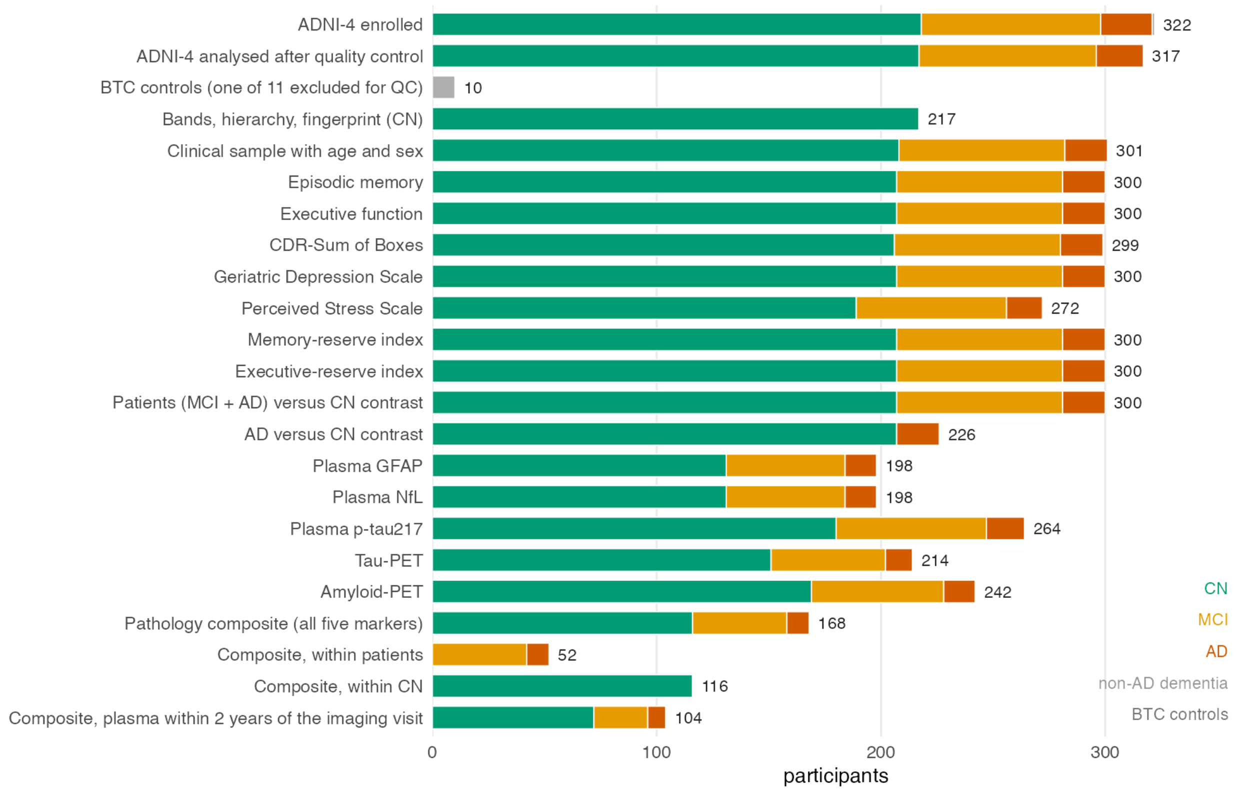


***Supplementary Figure 1. Participant inclusion and sample sizes across datasets and analyses.*** *The figure summarizes the number and diagnostic composition of participants included in each analysis. The ADNI-4 cohort comprised 322 enrolled participants, of whom 317 were retained after quality control and exclusion of one participant with non-AD dementia. Analyses requiring age and sex information included 301 participants. The BTC methodological dataset included 10 healthy controls; 11 controls were assessed, with one excluded during quality control. The number of participants available varied across clinical measures and analyses according to data availability. The pathology composite was calculated in 168 participants with complete data for all five biomarkers, including 116 cognitively normal (CN) participants and 52 participants with mild cognitive impairment (MCI) or Alzheimer's disease (AD). The final row shows the subset with plasma samples collected within two years of the imaging visit. Bar colours indicate diagnostic group; grey indicates BTC controls.*

### S2. Sensitivity of individual identifiability to the definition of short-range connectivity.

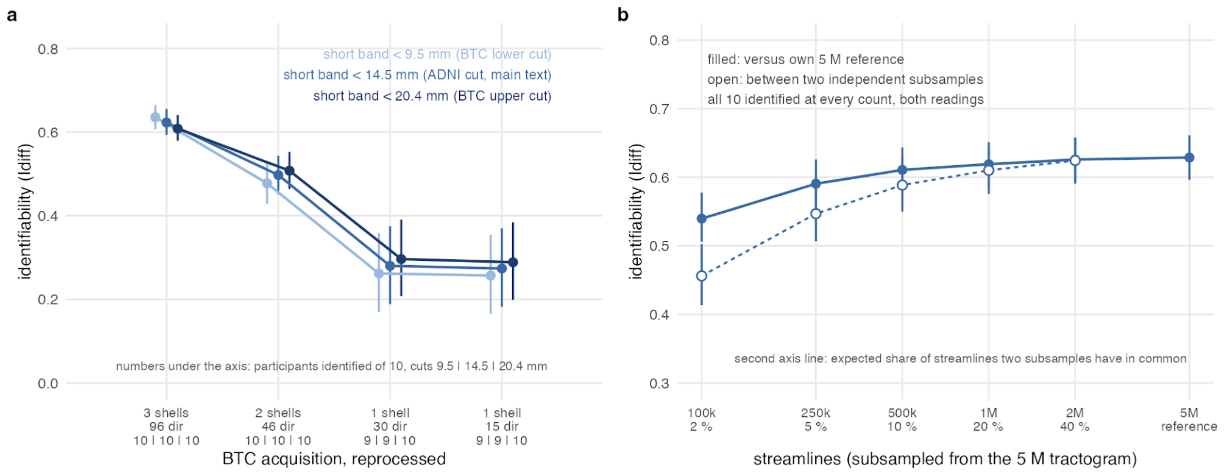


***Supplementary Figure 2. Sensitivity of individual identifiability to the definition of short-range connectivity.*** *Individual identifiability (IDIFF) was evaluated in the 10 BTC controls using three alternative definitions of short-range connectivity: connections shorter than 9.5 mm, 14.5 mm, or 20.4 mm. These thresholds correspond to the lower-tertile boundary in BTC, the threshold used in the main ADNI-4 analyses, and the upper-tertile boundary in BTC, respectively. Identifiability was evaluated across the original three-shell acquisition and progressively reduced single- and two-shell configurations. Numbers below the horizontal axis indicate the number of participants identified out of 10 for each threshold. Error bars indicate 95% confidence intervals from 10,000 participant-level bootstrap resamples.*

### S3. Comparison of connection-distance distributions and weighted-tertile boundaries between ADNI-4 and BTC.

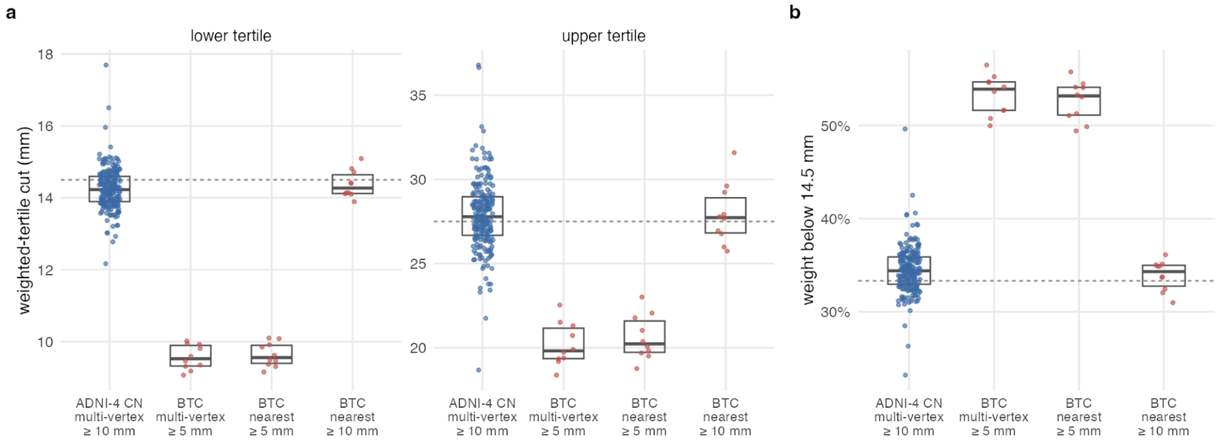


***Supplementary Figure 3. Comparison of connection-distance distributions and weighted-tertile boundaries between ADNI-4 and BTC****. (a) Distribution of the lower- and upper-tertile distance cutoffs across participants in ADNI-4 and BTC. Each point represents an individual participant; boxplots summarize the distribution within each dataset. Dashed horizontal lines indicate the corresponding cutoffs derived from the ADNI-4 cohort. (b) Proportion of total connection weight contributed by connections shorter than 14.5 mm in each participant. The dashed horizontal line indicates the mean proportion in the ADNI-4 cohort. BTC results are shown separately for the multi-vertex connectome and the nearest-vertex assignment, using minimum streamline lengths of 5 mm or 10 mm, as indicated.*

### S4. Individual identifiability across spatial scales and its relationship with cortical coverage.

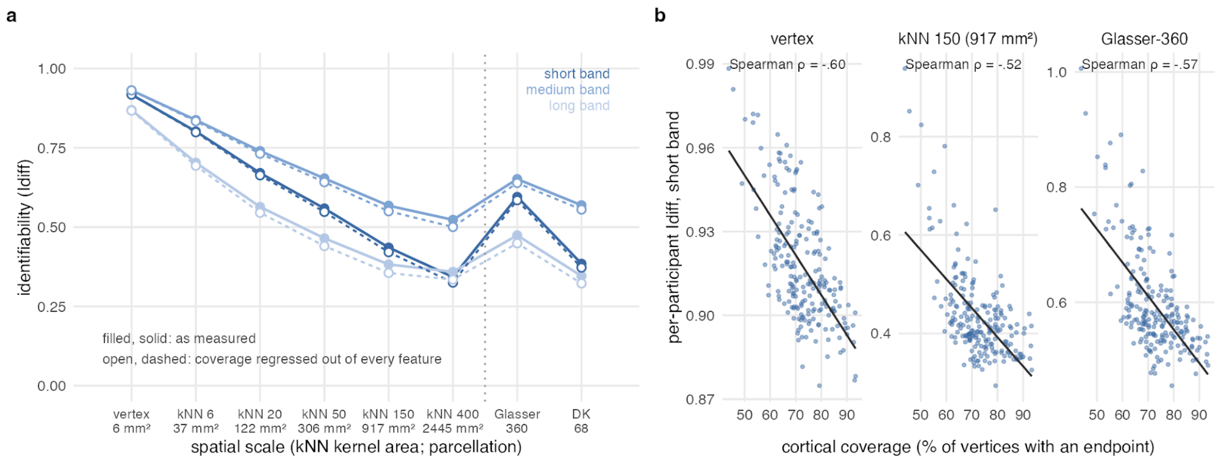


***Supplementary Figure 4. Individual identifiability across spatial scales and its relationship with cortical coverage.*** *(a) Individual identifiability (Idiff) for short-, medium-, and long-range connections across spatial scales, from the native vertex-wise connectome to progressively larger k-nearest-neighbour (kNN) smoothing kernels and atlas-based parcellations. Filled symbols and solid lines show the measured values; open symbols and dashed lines show values after regressing cortical coverage out of each connectivity feature. The vertical dotted line separates vertex-wise smoothing scales from atlas-based parcellations. (b) Relationship between individual identifiability of short-range connectivity and cortical coverage at the vertex-wise, kNN 150, and Glasser-360 scales. Each point represents a participant. Spearman correlation coefficients are indicated in each panel.*

### S5. Clinical associations and sensitivity of diagnostic contrasts to acquisition-site adjustment

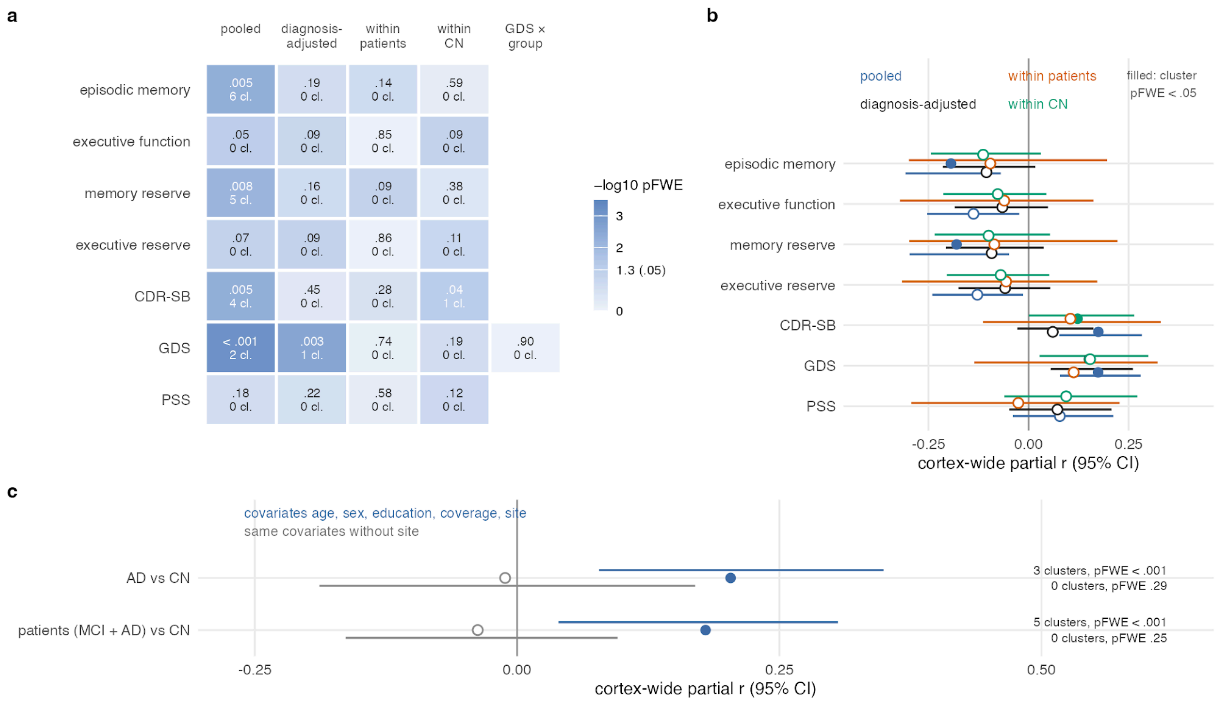


***Supplementary Figure 5. Clinical associations and sensitivity of diagnostic contrasts to acquisition-site adjustment.*** *(a) Summary of cluster-level associations between the short-to-long connectivity balance and clinical measures. Heatmap values indicate cluster-level family-wise error-corrected significance ($p_{FWE}$), with the number of significant clusters indicated in each cell. Analyses are shown for pooled participants, diagnosis-adjusted models, and analyses restricted to participants with MCI or AD or to cognitively normal (CN) participants. The GDS-by-diagnostic-group interaction is also shown. (b) Cortex-wide partial correlation coefficients and 95% confidence intervals for the clinical measures across the pooled, diagnosis-adjusted, within-patient, and within-CN analyses. Filled symbols indicate associations with at least one significant cluster ($p_{FWE}$ < .05). (c) Diagnostic contrasts comparing AD with CN participants and participants with MCI or AD with CN participants, with and without adjustment for acquisition site. Points indicate cortex-wide partial correlation coefficients and horizontal lines indicate 95% confidence intervals. The number of significant clusters and corresponding $p_{FWE}$ values are indicated for each model.*

#### S6. MRI acquisition protocols

The BTC reference cohort comprised the 10 healthy control participants of the BTC dataset (4 female; age 58.6 ± 10.3 years, range 48–75), scanned at Ghent University Hospital on a 3 T Siemens Magnetom Trio (TrioTim, syngo MR B17). The T1-weighted image used a 3D MPRAGE sequence at 1.0 mm isotropic resolution (matrix 256 × 256, 144 sagittal slices) with a repetition time of 1.59 s, an echo time of 4.18 ms, an inversion time of 900 ms, a flip angle of 9°, and twofold in-plane parallel acceleration. Diffusion-weighted images used a single-shot spin-echo echo-planar sequence at 2.5 mm isotropic resolution (matrix 96 × 96, 60 slices) with a repetition time of 8.7 s, an echo time of 110 ms, a flip angle of 90°, and twofold acceleration.

The multi-shell scheme comprised 6 non-diffusion-weighted (b = 0) volumes and 16, 30, and 50 directions at b = 700, 1200, and 2800 $s \cdot mm^{-2}$ (102 volumes, anterior-to-posterior phase encoding), with 2 further b = 0 volumes acquired with reversed phase encoding for susceptibility-distortion correction. The clinical cohort followed the standardized ADNI-4 MRI protocol. Data was acquired at 3 T (Siemens Magnetom Prisma / Prisma Fit) using harmonized sequences across participating sites; cross-vendor and software variation is described in the ADNI-4 protocol documentation (Weiner et al. 2017); ADNI MR Core).

The T1-weighted image was an accelerated three-dimensional MPRAGE acquisition at 1 mm isotropic resolution, consistent with the ADNI structural reference protocol. Diffusion-weighted images were acquired with a two-dimensional multi-band multi-shell echo-planar sequence, with a repetition time of 3400 ms, an echo time of 71 ms, and a flip angle of 90°. The diffusion scheme comprised 13 non-diffusion-weighted (b = 0) volumes together with 6 directions at b = 500 $s \cdot mm^{-2}$, 48 directions at b = 1000 $s \cdot mm^{-2}$, and 60 directions at b = 2000 $s \cdot mm^{-2}$, for a total of 127 measurements.

| **Parameter** | **Reference cohort ($BTC_{preop}$)** | **Clinical cohort (ADNI-4)** |
|---|---|---|
| Site / scanner | Ghent University Hospital; 3 T Siemens Magnetom Trio | Multi-site; 3 T Siemens Magnetom Prisma / Prisma Fit |
| **T1-weighted** | | |
| Sequence | 3D MPRAGE | Accelerated 3D MPRAGE |
| Resolution | 1 mm isotropic (matrix 256) | 1 mm isotropic (208 slices, matrix 256 × 240) |
| TR / TE / TI | 1.59 s / 4.18 ms / 900 ms | 2.3 s / 2.96 ms / 900 ms |
| Flip angle | 9° | 9° |

**Diffusion-weighted**

| | | |
|---|---|---|
| Sequence | Single-shot spin-echo EPI | 2D multi-band multi-shell EPI |
| Resolution | 2.5 mm isotropic | 2.0 mm isotropic |
| TR / TE | 8.7 s / 110 ms | 3.4 s / 71 ms |
| Flip angle | 90° | 90° |
| Shells (directions) | b = 700 (16), b = 1200 (30), b = 2800 (50) $s\cdot mm^{-2}$ | b = 500 (6), b = 1000 (48), b = 2000 (60) $s\cdot mm^{-2}$ |
| b = 0 volumes | 6 (AP) + reverse-PE (PA) | 13 |
| Total volumes | 102 (AP series) | 127 |

***Supplementary Table 1. MRI acquisition parameters of the reference (BTC) and clinical (ADNI-4) cohorts***.

ADNI-4 acquires diffusion data under a two-tier scheme: an Advanced multi-shell protocol on multiband-capable scanners and a Basic single-shell protocol elsewhere. Because the degradation analysis identified multi-shell sampling as the binding requirement for recovering subject-specific structure, only participants scanned with the Advanced protocol were included; the parameters above describe that acquisition.
